# Co-occurrence of Seizures and Gastrointestinal Symptoms in Children with ASD

**DOI:** 10.1101/2025.02.25.25322888

**Authors:** Alessandro Tagliavia, Andrey Vyshedskiy

**Affiliations:** Boston University, Boston MA 02215, USA

## Abstract

The goal of this cluster analysis of comorbidities in children with Autism Spectrum Disorder (ASD) was to find potential co-occurrence between health symptoms that would not otherwise be investigated. We analyzed parent-reported data from 130,985 users of a free language-learning application. The initial cohort was stratified by ASD severity and age group to explore patterns within these subpopulations. Across these cohorts and subgroups, we observed a consistent co-occurrence of gastrointestinal symptoms, such as diarrhea and constipation, with seizures and lethargy. These results suggest that a common underlying mechanism may be driving both gastrointestinal disturbances and seizure activity in children with ASD.

## Introduction

Autism Spectrum Disorder (ASD) is a neurodevelopmental condition characterized by a diverse array of symptoms and comorbidities affecting multiple body systems. One system that has been often studied in relation to ASD has been the gastrointestinal (GI) system. A systematic review of thirty articles found that children with ASD are significantly more likely to experience GI symptoms than healthy controls (Leader et al., 2022), with some studies noting that GI symptoms are four times as prevalent in children with ASD, with diarrhea and constipation being the most common (Saurman et al., 2020). This has become an important part of ASD research in recent years for a variety of reasons. GI symptoms can aggravate existing behavioral symptoms (Socała et al., 2021). For example, Ferguson et al found that in younger children, constipation was linked to greater rates of anxiety, while nausea was linked with aggressive behaviors (Ferguson et al., 2019). In older children, stomachaches were linked to increased withdrawn behavior (Ferguson et al., 2019). There is also the potential for establishing a link between GI factors, such as intestinal microbiota, and neurodevelopment of children with ASD (Wasilewska et al., 2015; Rao et al., 2016). It has also been reported that children with ASD are at greater risk of epilepsy and seizures (Liu et al., 2022), and that GI disorders in children with ASD can increase the risk of epilepsy (Al-Beltagi et al., 2023). In a 2015 retrospective study, Aldinger et al reviewed the medical information of roughly 3,000 individuals across two cohorts to explore the connection between GI symptoms and ASD. The authors found a significant relationship between GI symptoms and seizures, and concluded that children with ASD who experience GI distress should also be screened for seizures (Aldinger et al., 2015).

In this study, we report our findings of consistent correlation between seizures and GI symptoms such as constipation and diarrhea, as well as lethargy as reported by caregivers via an app. In 2015 we developed a language therapy app and made it available gratis at all major app stores (Vyshedskiy et al., 2015; Vyshedskiy et al., 2020; Dunn et al., 2017a; Dunn et al., 2017b; Dunn et al., 2017c). The app is primarily popular with autistic children and therefore the majority of our participants are young children diagnosed with ASD. In return for access to structured language exercises inside the app, parents are asked to complete a 133-question survey every three months.

This study looks at cluster analyses of children by age, as well as by severity of ASD, to explore potential associations between GI symptoms and seizures in children with ASD.

## Methods

### Participant Selection

Participants were users of a language therapy app that was made available gratis at all major app stores in September 2015. Once the app was downloaded, caregivers were asked to register and provide demographic details, including the child’s diagnosis and age. Caregivers consented to anonymized data analysis and completed the Autism Treatment Evaluation Checklist (ATEC) (Rimland et al. 1999) and Mental Synthesis Evaluation Checklist (MSEC) (Arnold et al., 2022; Braverman et al. 2018) parent reports. The first evaluation was administered approximately one month after the app download. The subsequent evaluations were administered at approximately three-month intervals.

From this data, 23 variables of interest were selected and a cluster-analysis was performed in R to find which of these variables cooccurred with greater frequency than others. An unsupervised hierarchical clustering of children 2 to 22 years of age was conducted, and once clusters were identified, the data was further analyzed with Principal Component Analysis (PCA) to see how consistently these variables co-occurred.

Because this application is often used by the same individuals over the course of several years, the data was cleaned to ensure that there was only one assessment per user used in the study. After this filtering, 131,721 users remained. Users were first grouped by ASD severity diagnosed by clinicians and reported by parents: Mild/Level I (n=59,974), Moderate/Level II (n=34,081), or Severe/Level III autism (n=37,666). The users were then grouped by age, analyzing each age group from 2 to 22 years of age. The number of participants by age is displayed in Table 1. PCA and unsupervised hierarchical cluster analyses were performed on these groups, and displayed in dendrograms.

**Table 1.** Participant age groups by year.

| Number of Participants by Age |  |  |  |
| --- | --- | --- | --- |
| 2-year-olds | 18,594 | 8-year-olds | 5,213 |
| 3-year-olds | 29,666 | 9-year-olds | 3,500 |
| 4-year-olds | 26,217 | 10-year-olds | 2,445 |
| 5-year-olds | 18,006 | 11-year-olds | 1,812 |
| 6-year-olds | 12,246 | 12-year-olds | 1,342 |
| 7-year-olds | 8,435 | 13–22-year-olds | 4,245 |
| Total |  | 131,721 |  |

### Statistical Method

Cluster analyses are useful statistical tools that allow the detection of the item co-occurrence that may not be immediately apparent otherwise. For example, if researchers had a database that included a population’s health information, and wanted to find what genes may predispose someone for a particular disease, a cluster analysis could group the genes that co-occur in individuals with a particular disease. If Genes A, B, and C, consistently co-occurred in individuals with heart disease, but Genes X, Y, and Z did not, researchers could reasonably hypothesize that Genes A, B and C either impact, or are impacted by, heart disease.

Similarly, we conducted a cluster analysis to group symptoms experienced by individuals with ASD. It is possible that the groupings found in cluster analyses can find previously unknown or unconfirmed correlations, and direct future study to explore the strength and direction of these links.

Unsupervised Hierarchical Cluster Analysis (UHCA) was performed using Ward’s agglomeration method with a Euclidean distance metric. Clustering analysis was data-driven without any design or hypothesis. PCA was also performed on the data to further validate the clusters identified by the unsupervised hierarchical clustering analysis. Code is available from the corresponding author upon reasonable request.

## Results

The unsupervised hierarchical clustering analysis, which encompassed all levels of ASD severity in individuals aged 2 to 22, revealed a cluster of constipation, lethargy, diarrhea, and seizures (Figure 1). When hierarchical clustering was stratified by autism severity, the same four items appeared clustered together in each severity group (Mild, Moderate, and Severe, Figures S1-S6)

**Figure 1.**
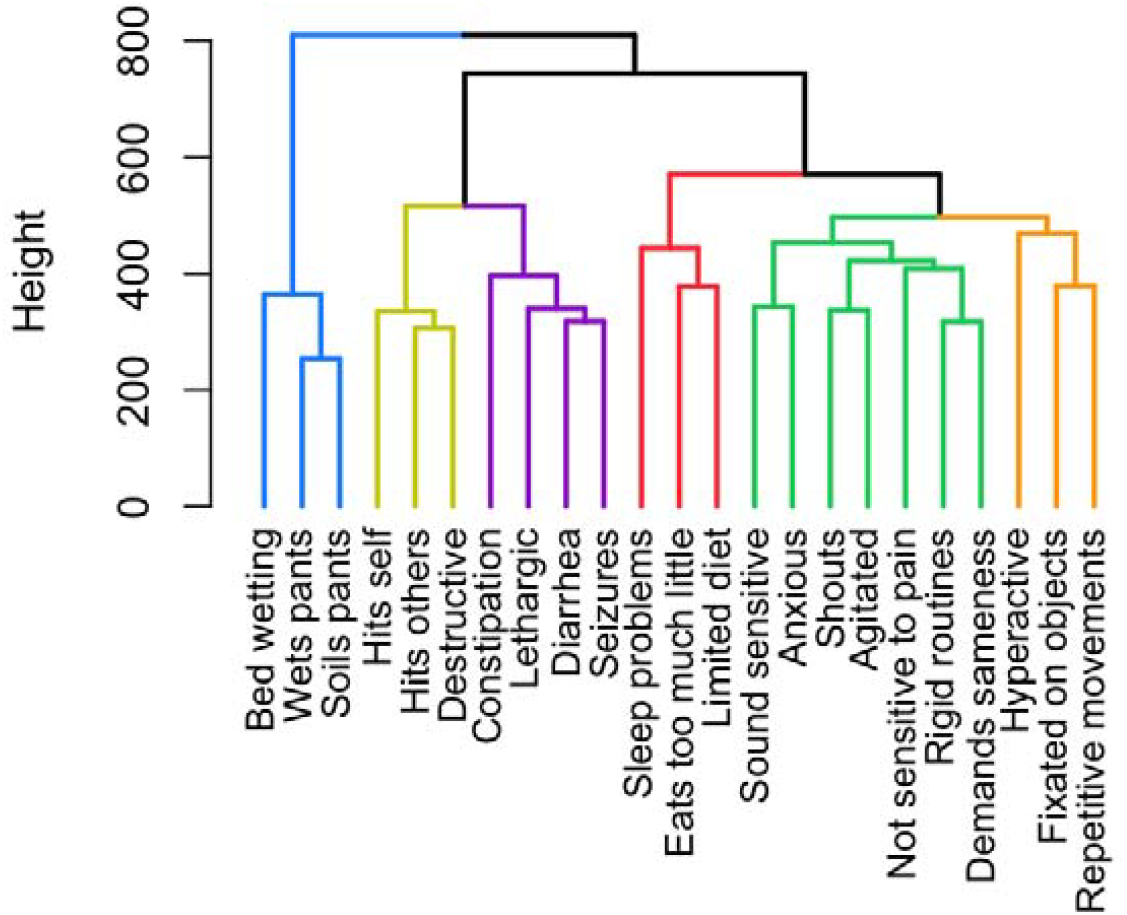
Dendrogram for unsupervised hierarchical analysis for ages 2-22 of all autism severities

For the 2-to 22-year-old group, PCA analysis found that PC1 (16.4%) and PC2 (15.8%) explain 32.3% of the total variance. The variables were tightly clustered, while constipation, lethargic, diarrhea, and seizures formed a distinct subgroup (Figure 2). Other variables that formed a distinct subgroup were soils pants, wets pants, and bed wetting.

**Figure 2.**
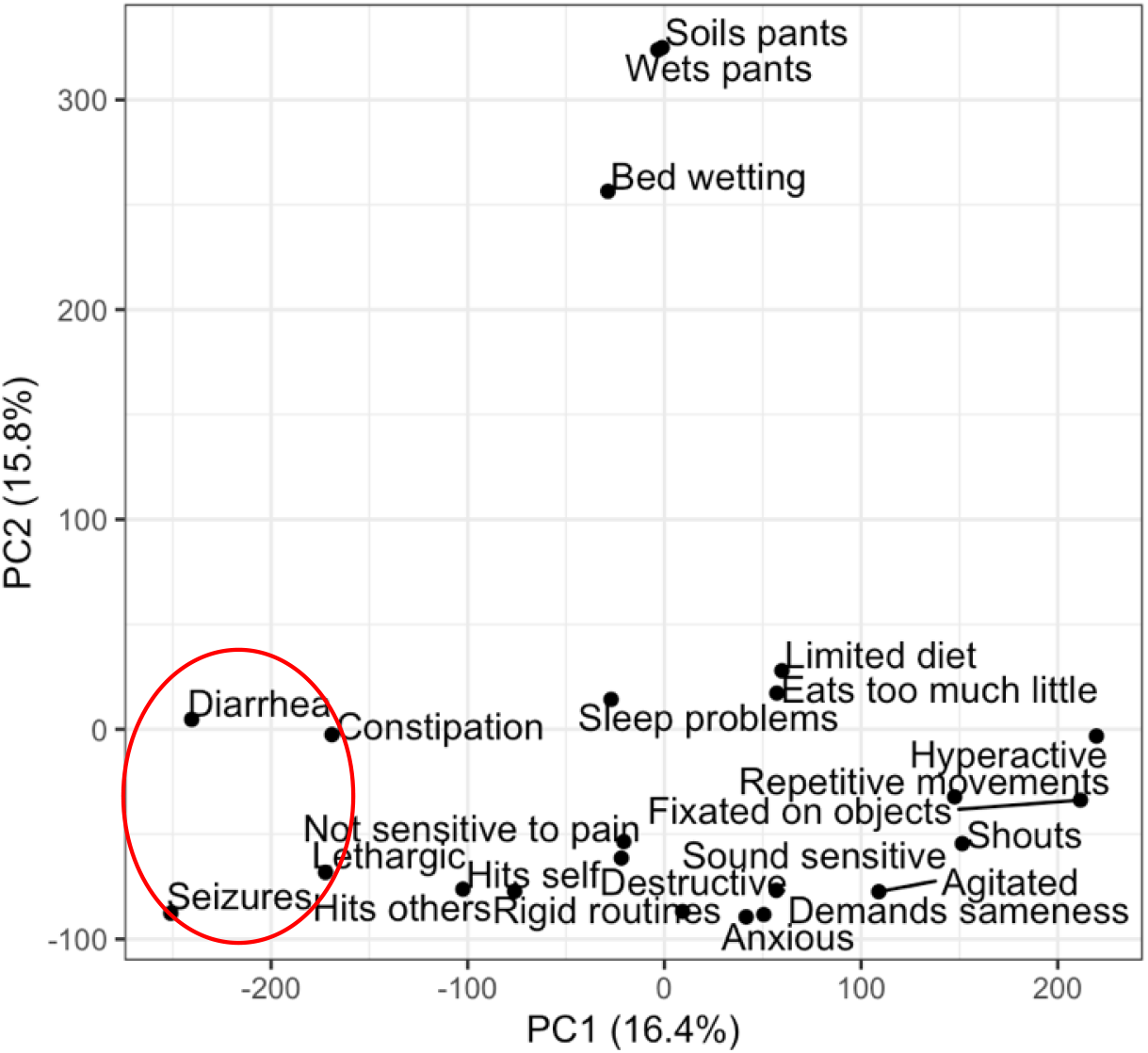
Principal Component Analysis for ages 2-22 of all autism severities

PCAs for each ASD severity groups showed similar findings. In each, the four items of interest clustered together, however the distance between the clusters was most distinct in the PCA for the Severe Autism subgroup.

Clustering individual age groups showed similar results to the overall dataset. Dendrograms and PCA analyses were conducted for each age group that had at least 1,000 participants. As our study population was predominately made up of younger children, this encompassed each whole age between 2-12 years. The dendrograms for these younger age groups were largely similar, with the variables of constipation, diarrhea, lethargy, and seizures, always appearing in the same cluster. For the older participants, we combined the remaining ages. An analysis of all of the older participants (age 13-22) combined showed clustering consistent with the younger age groups.

Which symptoms were closer to each other within the clusters also varied. For the younger participants, diarrhea and seizures were clustered in the lowest branch when not separated by age. Once age is accounted for, this finding is conserved in only six of the eleven age groups (4-, 5-, 6-, 7-, 8-, and 9-year-olds). Three of the remaining age groups showed seizures most closely related to lethargy (2-, 3-, and 12-year-olds), while the 10 and 11-year-old groups showed seizures closely related to the diarrhea and constipation symptoms.

The analysis of older participants (age 13-22), saw seizures most closely related to diarrhea, which is consistent with the findings for the unsupervised clustering of all age- and severity-groups in Figure 1.

PCA conducted by age group displayed a distinct clustering of the four variables of interest. For ages 2-4, this cluster was found in the top right quadrant (Figures S19-21), and most closely resembled the PCA of the Severe Autism subset (Figure S6).

For the remaining PCAs, the cluster was mostly conserved, though appearing in the bottom left quadrant (Figures S22-30). Beginning with the 7-year-old subgroup, the other variables become less tightly clustered, though there is still clear grouping of the four variables of interest (Figures S24-30).

The differences in variance by PC1 and PC2 are provided in Table S1.

## Discussion

While a link between ASD and GI symptoms has been observed in the past (Leader et al., 2021; Saurman et al., 2020; Socała et al., 2021; Ferguson et al., 2019; Wasilewska et al., 2015; rao et al., 2016; Srikantha et al., 2019; Wang et al., 2023), our access to a large dataset of children with ASD and their parent-reported symptoms offers an ability to cluster specific symptoms at a large scale. Studying a dataset of this size allows better detection of co-occurring comorbidities. Clustering arranges the symptoms by their co-occurrence. This means that if two comorbidities are mediated by the same mechanism, and this mechanism is introduced or removed, the clustered comorbidities should be affected with similar timing and magnitude. Importantly, cluster analyses perform grouping calculations in a data-driven manner without necessitating a hypothesis, allowing co-occurring comorbidities to be identified without preexisting knowledge of their linkage (Zhang et al., 2017; Dalmaijer et al., 2022).

From this large dataset we were able to see links between particular comorbidities. Two groups of comorbidities were shown to be distinct from the others. One, the triad of soils pants, wets pants, and bed-wetting, were of little surprise. However, the consistent grouping of diarrhea, constipation, seizures, and lethargy, particularly the frequent subgrouping of diarrhea and seizures, stood out.

The co-occurrence of GI symptoms and seizures in individuals with ASD was previously reported in a variety of studies (Saurman et al., 2020; Doshi-Velez et al., 2014; Mouridsen et al., 1999), with the co-occurrence of ASD and seizures being reported as far back as the 1970s (Mouridsen et al., 1999). However, the exact mechanism is unclear.

A variety of pathways and causes have been suggested for the occurrence of GI symptoms in children with ASD, as well as how those may link to seizures. These include a multitude of genetic and environmental factors. Several previous studies have looked at maternal immune activation (Rao et al., 2016; Wang et al., 2023; Hsiao et al., 2013; Zheng et al., 2021). One study in mice examined the offspring of mothers whose immune systems were activated via viral injection. The researchers found that these offspring exhibited several key symptoms of ASD observed in humans (Malkova et al., 2012).

Another common candidate is the leaky-gut hypothesis, which suggests that compromised intestinal permeability can lead to unwanted substances leaking into the bloodstream, and potentially having downstream impacts on signaling and other mechanisms (Saurman et al., 2020; Wasilewska et al., 2015; Rao et al., 2016; Srikantha et al., 2019; Wang et al., 2023). While there has been some evidence that certain restrictive diets can ameliorate ASD symptoms, the data is limited and not conclusive and requires further investigation.

When looking at seizures specifically, one study found a difference in gut microbiota between treatment-resistant epileptic patients and those who were treatment-responsive, with the latter group having a microbiota more similar to healthy controls (Peng et al., 2018). This suggests that gut dysbiosis, autism, and seizures, are all potentially linked along the gut-brain axis, and further investigation could help find a way to ameliorate symptoms of each of these.

Further, multiple possible mechanisms have been suggested to explain the co-occurrence of seizures and GI symptoms. A 2021 article by Beck et al. noted that potential mechanisms can include: decreased mobility leading to constipation, anti-seizure medications with side-effects that cause GI symptoms, enteral nutrition and the ketogenic diet, which are used to help with epileptic symptoms, leading to GI symptoms, as well as genetic variants in voltage-gated ion channels (Beck et al., 2021). Beck et al. also note that these same mechanisms can be present in individuals with ASD (Beck et al., 2021).

The gut-brain axis has been an area of interest in recent years, particularly as it interacts with ASD (Saurman et al., 2022; Socała et al., 2021; Rao et al., 2016; Liu et al., 2022; Srikantha et al., 2019; Wang et al., 2023; Beck et al., 2021; Cryan et al., 2019). Our hope is that reporting correlations between specific comorbidities along this gut-brain axis, such as the clustering of seizures and lethargy with diarrhea and constipation, can help future research as it looks to establishing a causal path for these comorbidities, as well as the conditions they cooccur with, particularly ASD and epilepsy.

## Limitations

This study utilizes exclusively parent reports, which can run the risk of not being as reliable as a health care provider’s report. However, parents interact with their children for several hours a day every day of the year, providing them with a deep knowledge of their child’s symptoms, particularly those involving gastrointestinal issues. Further, previous analyses of our own database support the reliability of parent reports (Fridberg et al., 2021; Jagadeesan et al., 2022).

The nature of large-scale datasets can also obscure the specific circumstances of a particular child’s symptoms. Yet it is this very size that allows us to find correlations that would not be obvious through case studies and anecdotes.

## Acknowledgements

We wish to thank all participants’ caregivers who found time to complete children’s assessments. The language therapy app used to collect the data presented in this manuscript was made possible by the contributions of Rita Dunn, Alexander Faisman, Jonah Elgart, Lisa Lokshina, and Yulia Dumov.

## Author contributions

AV and AT designed the study. AT wrote the paper.

## Informed Consent

Caregivers have consented to anonymized data analysis and publication of the results. The study was conducted in compliance with the Declaration of Helsinki ^85^.

## Compliance with Ethical Standards

Using the Department of Health and Human Services regulations found at 45 CFR 46.101(b)(4), the Biomedical Research Alliance of New York LLC Institutional Review Board (IRB) determined that this research project is exempt from IRB oversight.

## Data Availability

De-identified raw data from this manuscript are available from the corresponding author upon reasonable request.

## Code availability statement

Code is available from the corresponding author upon reasonable request.

