## Supplemental material for "Co-occurrence of Seizures and Gastrointestinal Symptoms in Children with ASD"

**Figure S1 – Mild Autism Dendrogram**

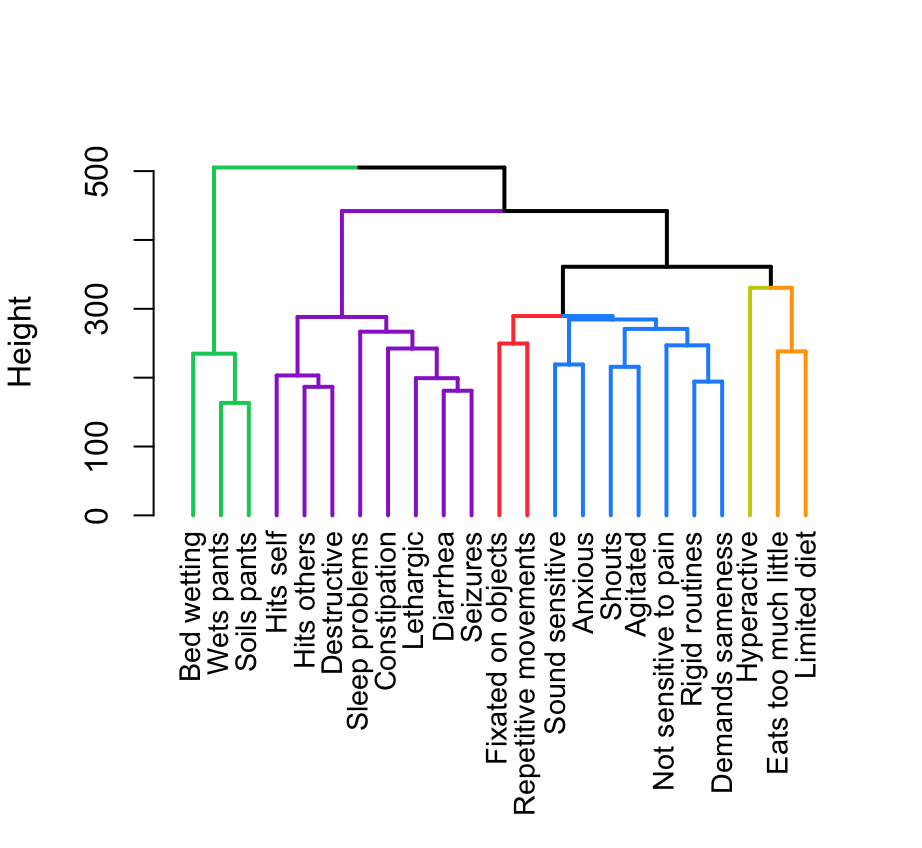

**Figure S2 – Moderate Autism Dendrogram**

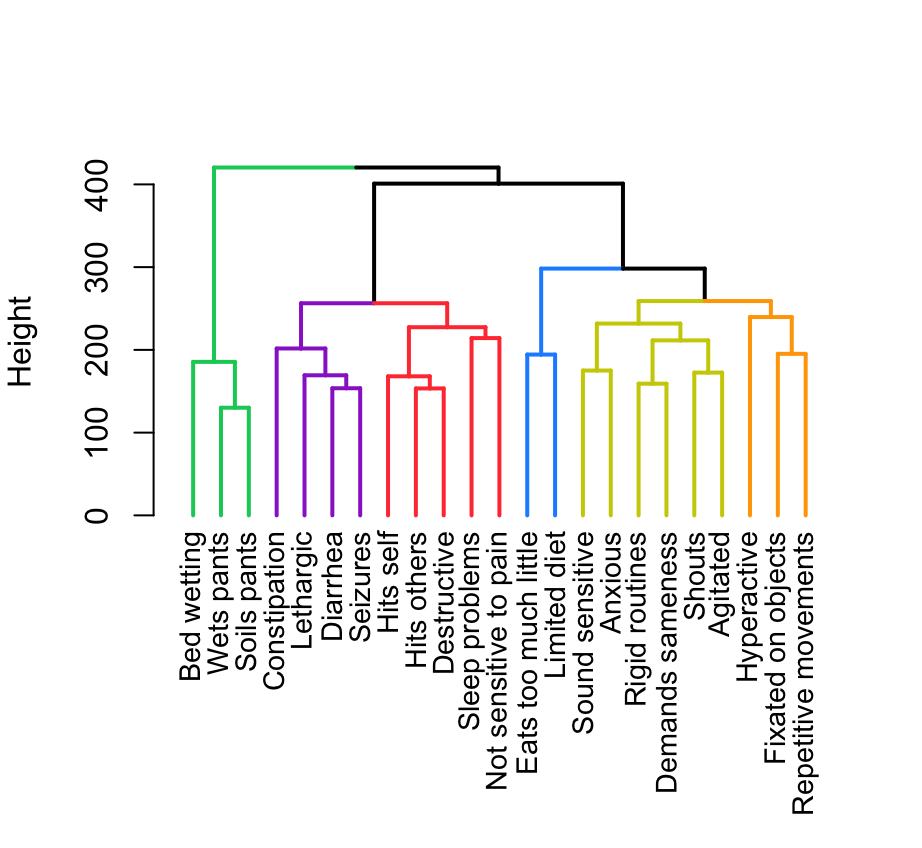

**Figure S3 – Severe Autism Dendrogram**

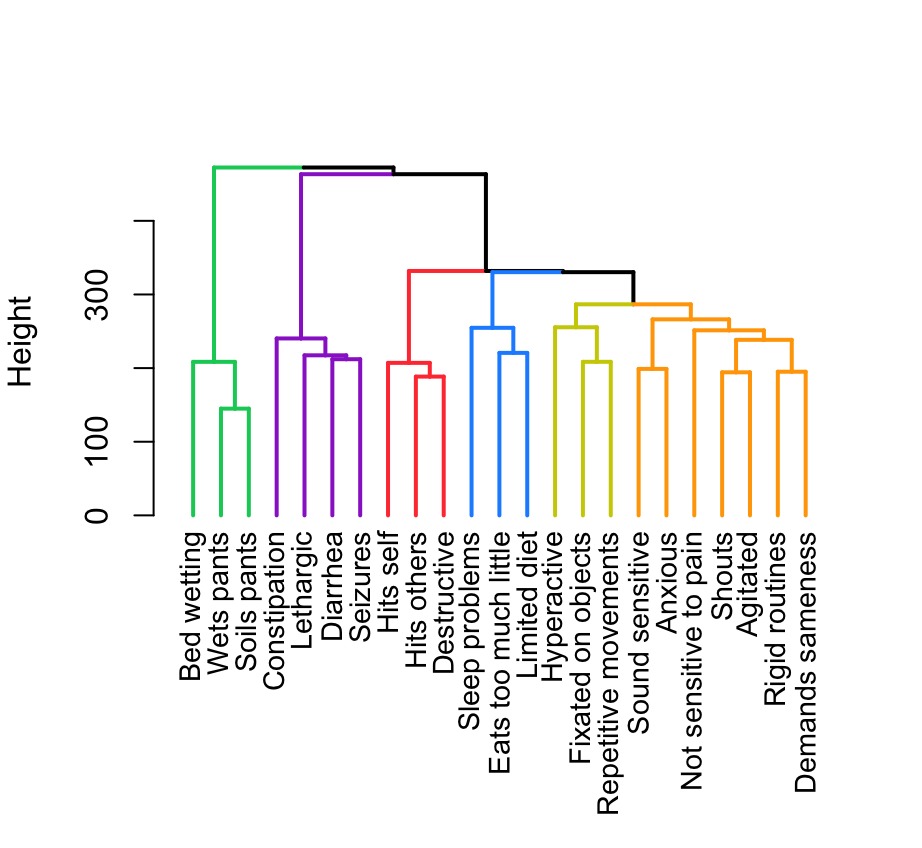

**Figure S4 – Mild Autism PCA**

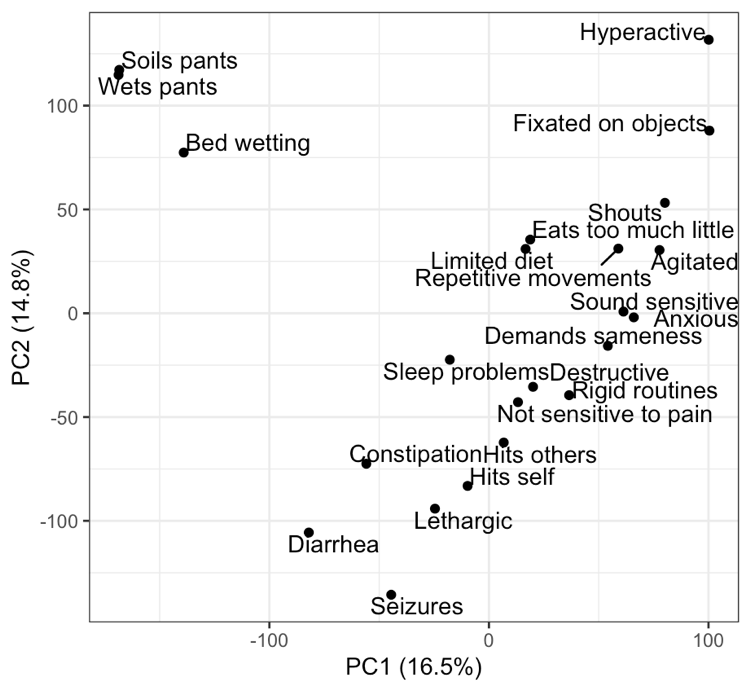

**Figure S5 – Moderate Autism PCA**

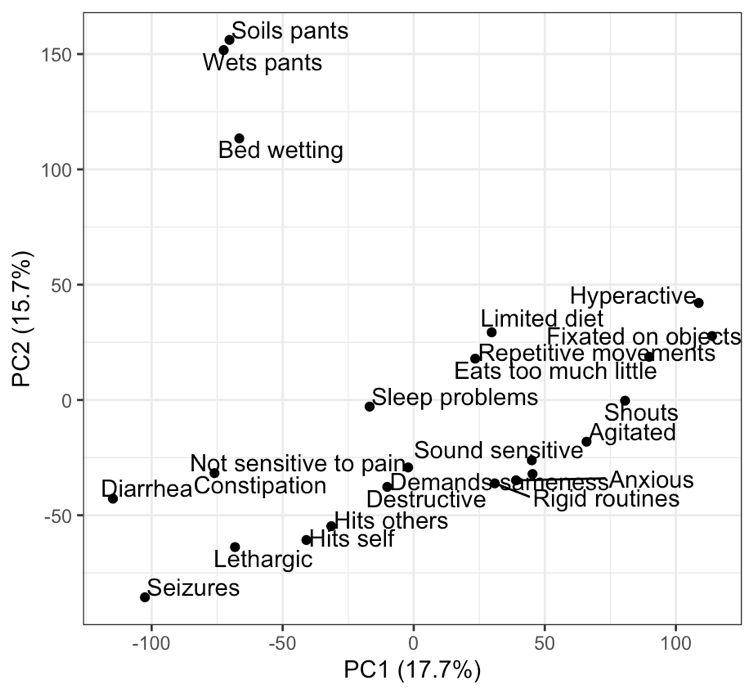

**Figure S6 – Severe Autism PCA**

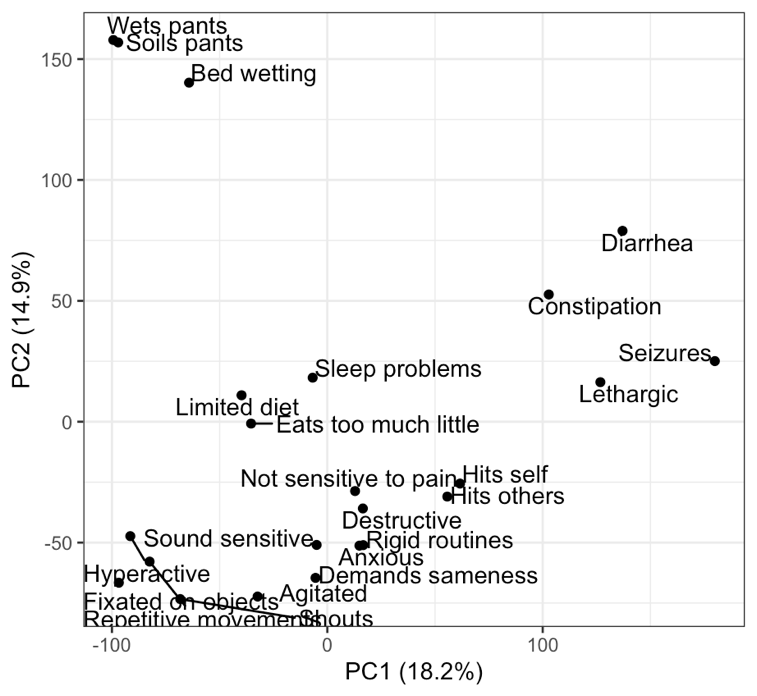

**Figure S7 - Age 2 Dendrogram**

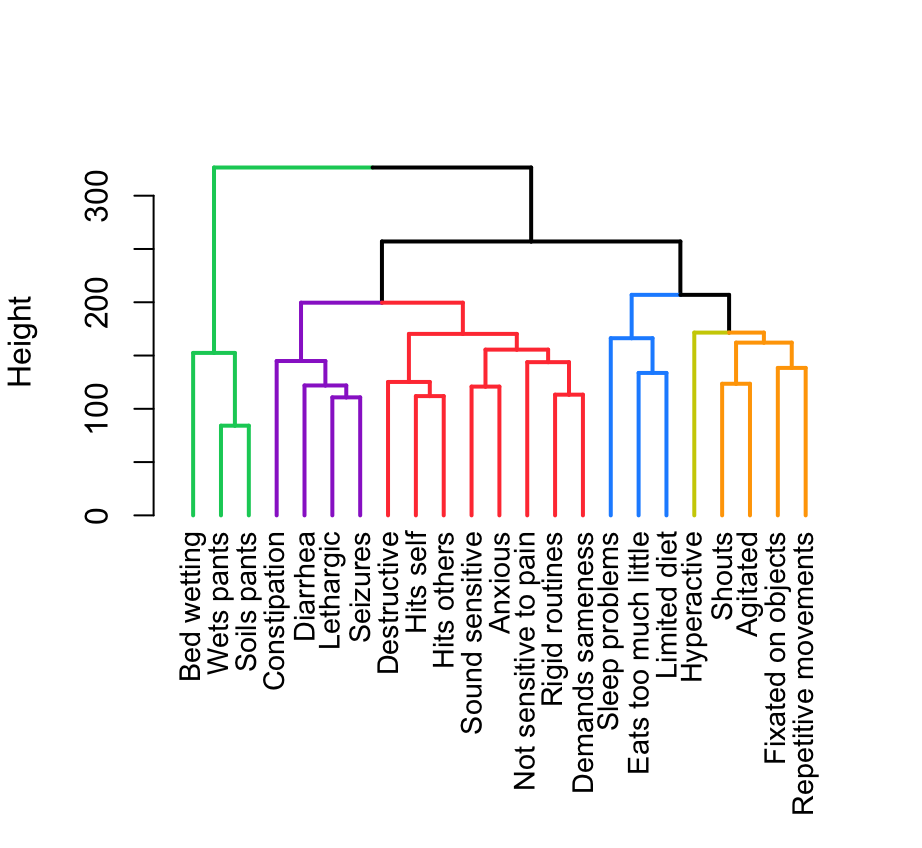

**Figure S8 - Age 3 Dendrogram**

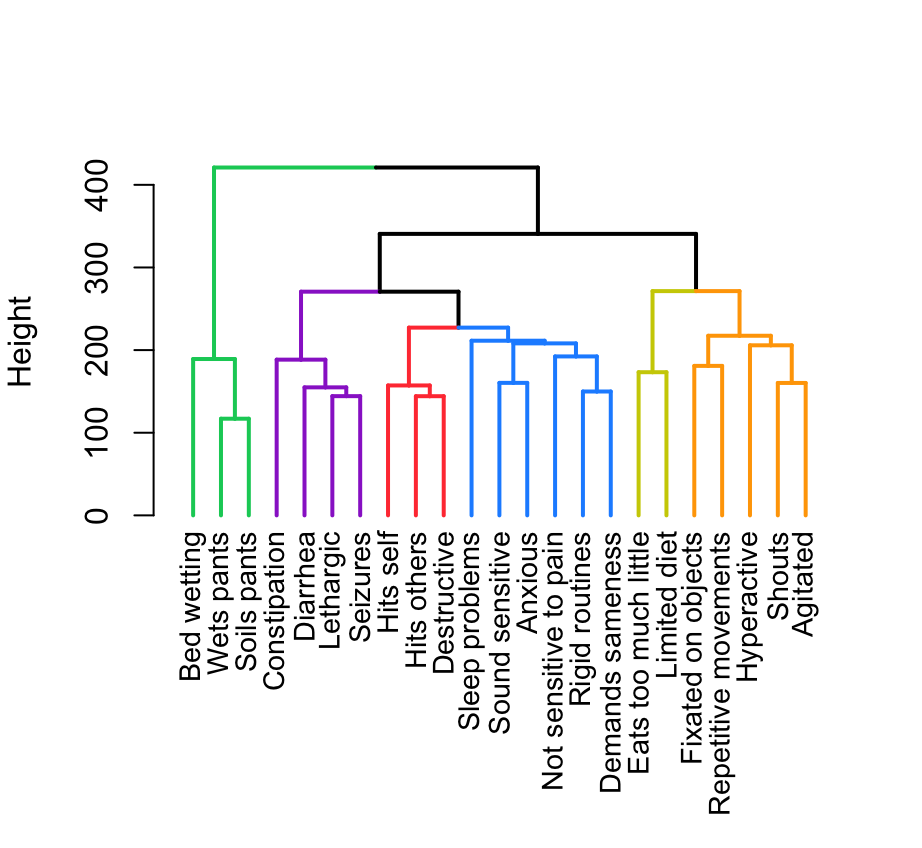

**Figure S9 - Age 4 Dendrogram**

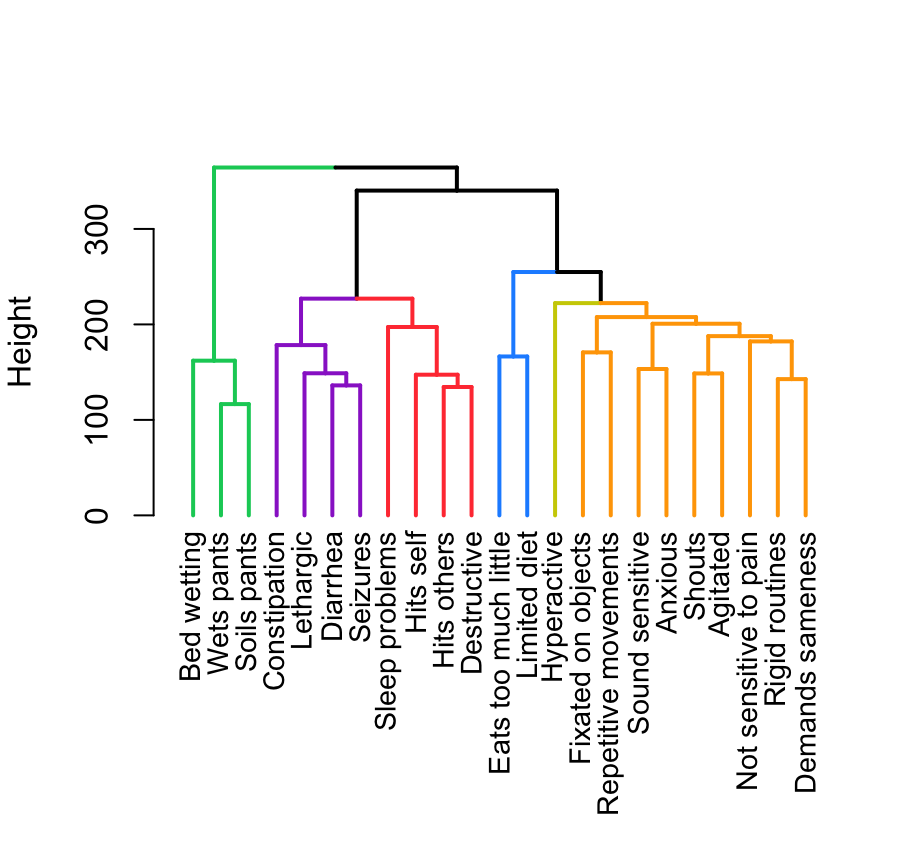

**Figure S10 - Age 5 Dendrogram**

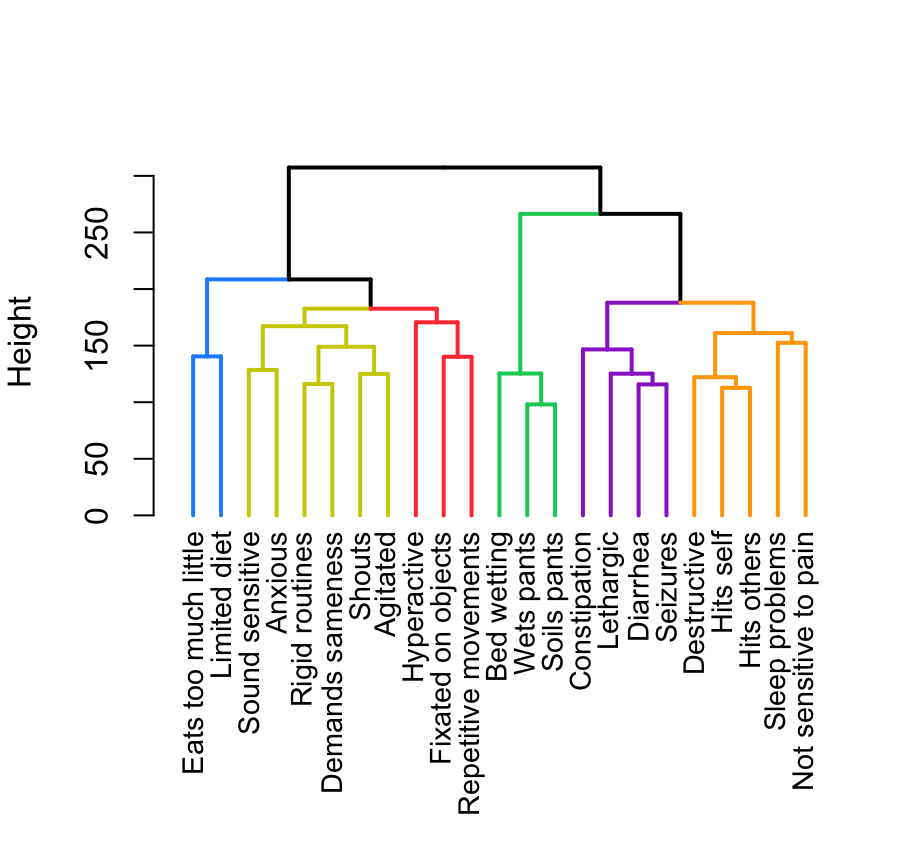

**Figure S11 - Age 6 Dendrogram**

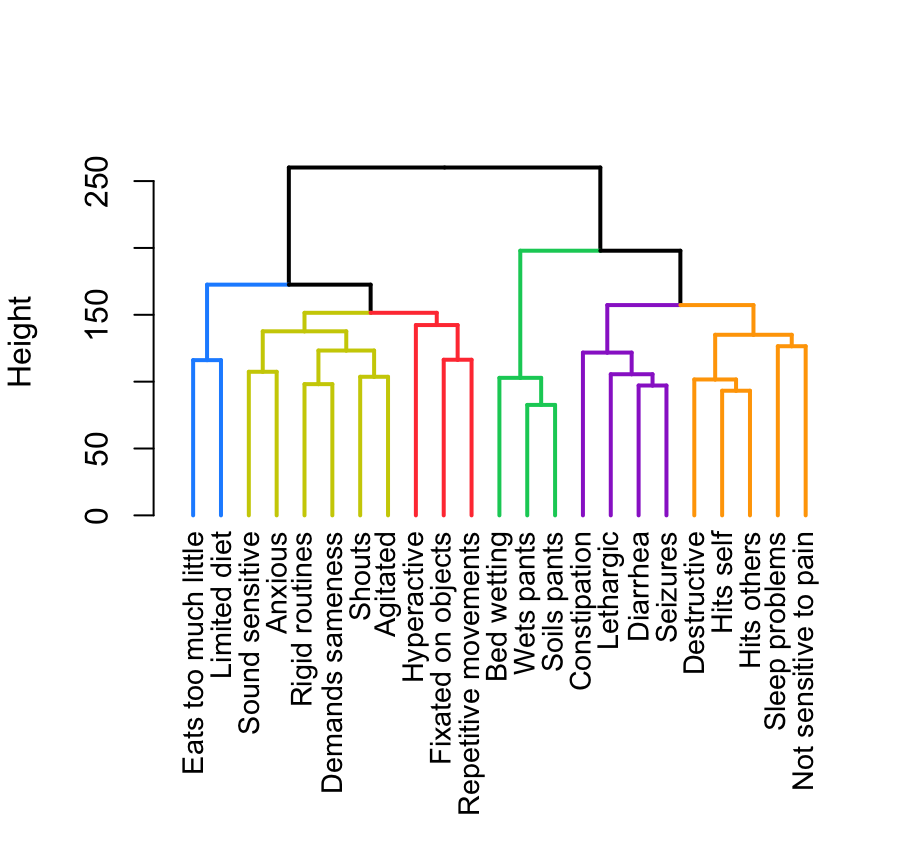

**Figure S12 - Age 7 Dendrogram**

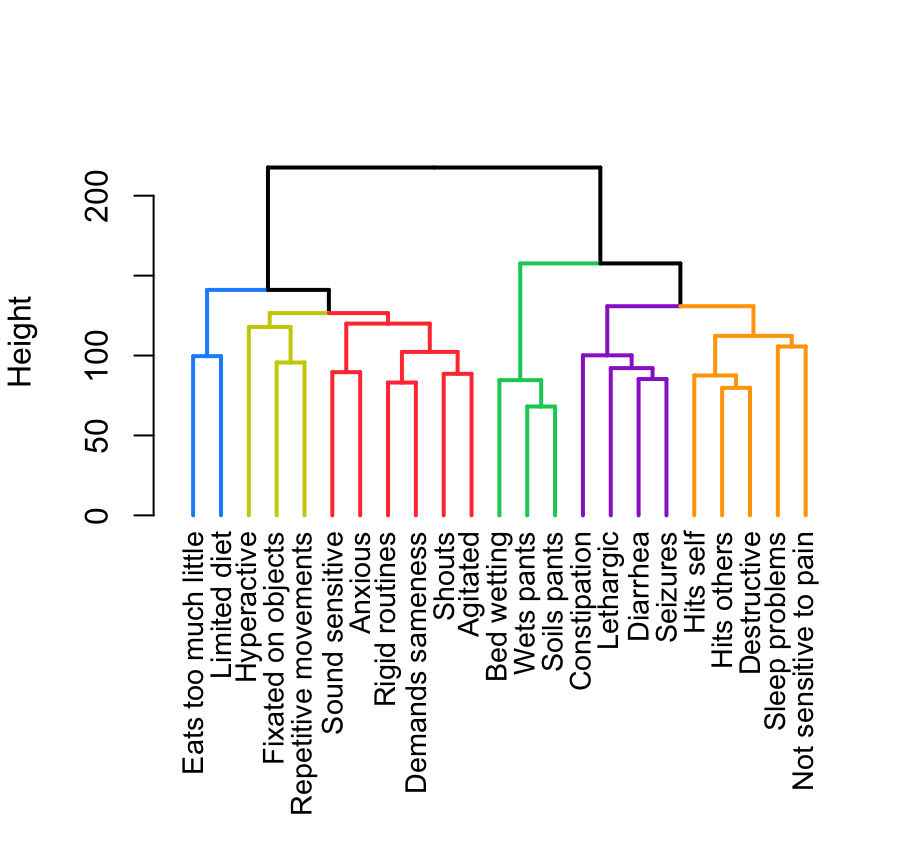

**Figure S13 - Age 8 Dendrogram**

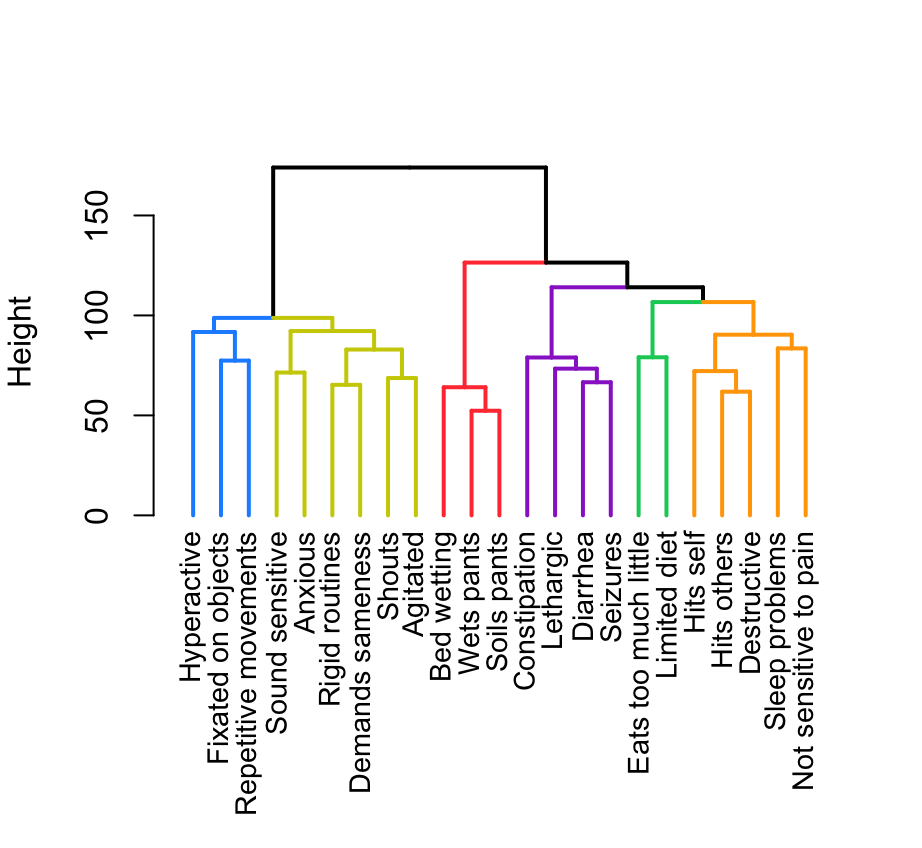

**Figure S14 - Age 9 Dendrogram**

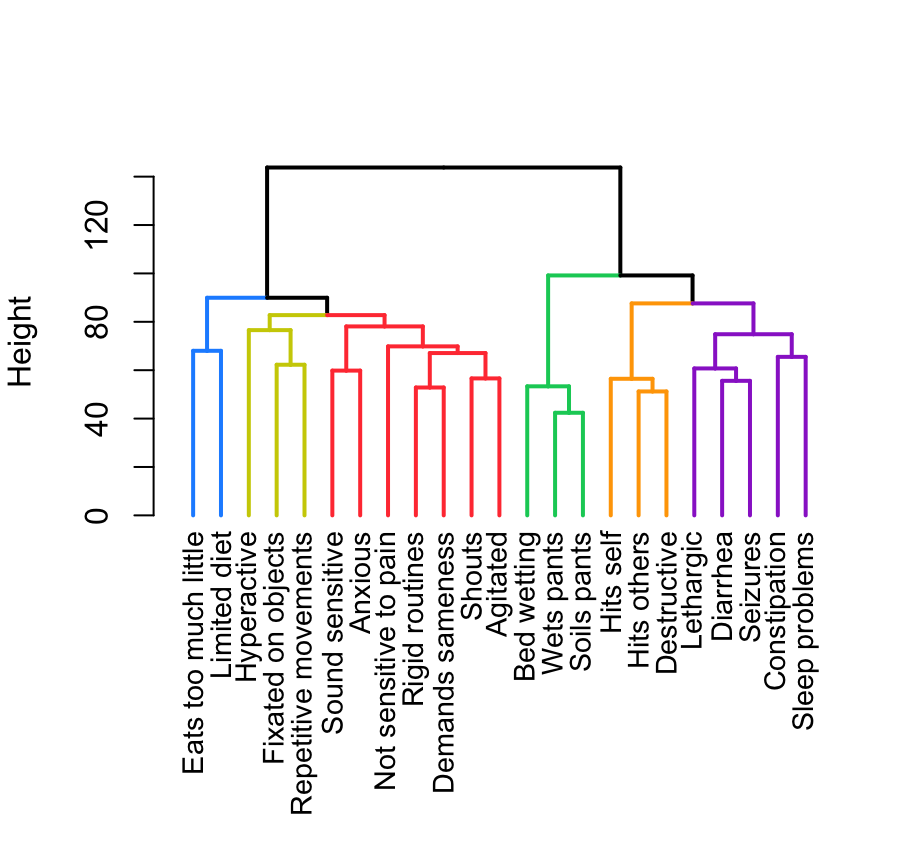

**Figure S15 - Age 10 Dendrogram**

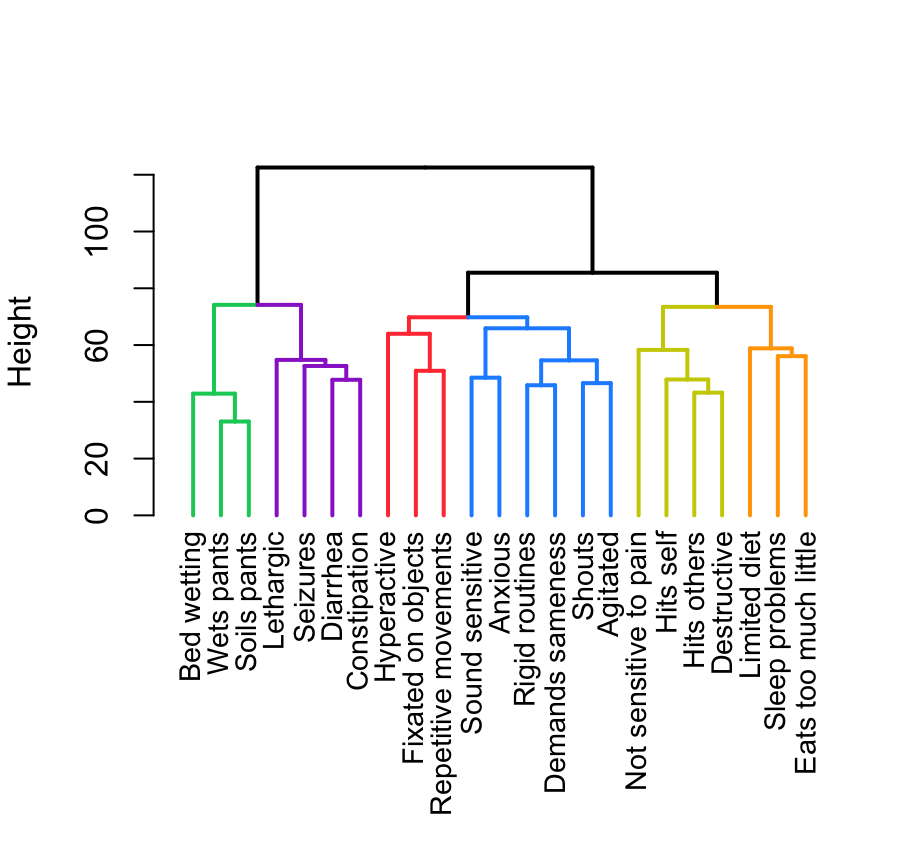

**Figure S16 - Age 11 Dendrogram**

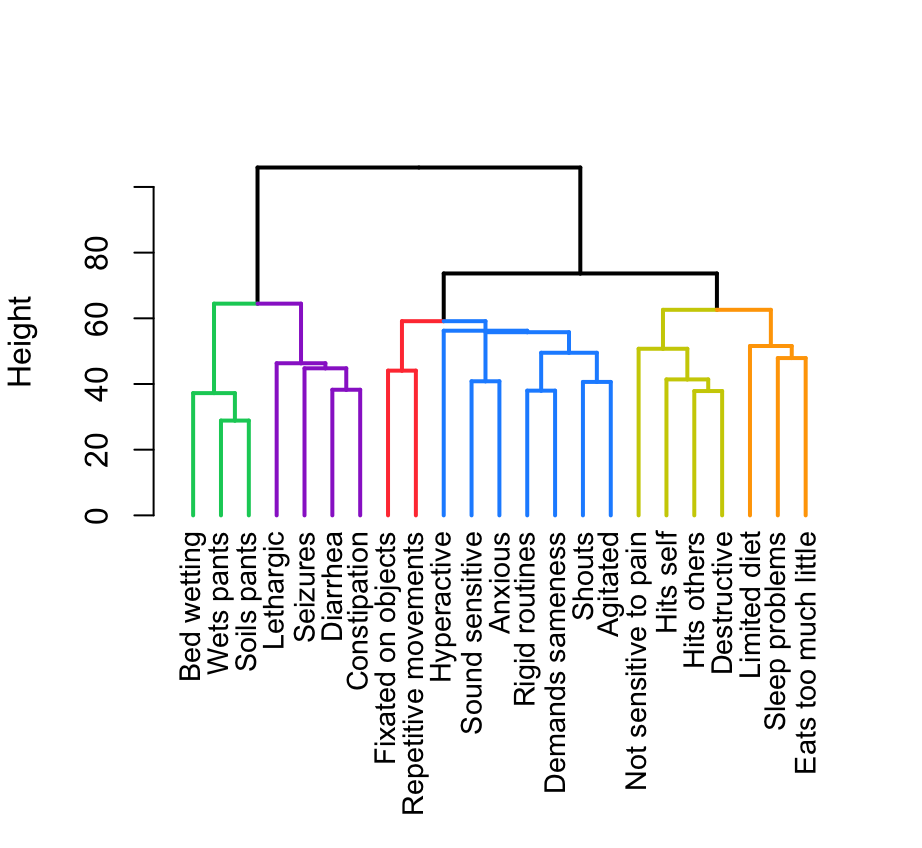

**Figure S17 - Age 12 Dendrogram**

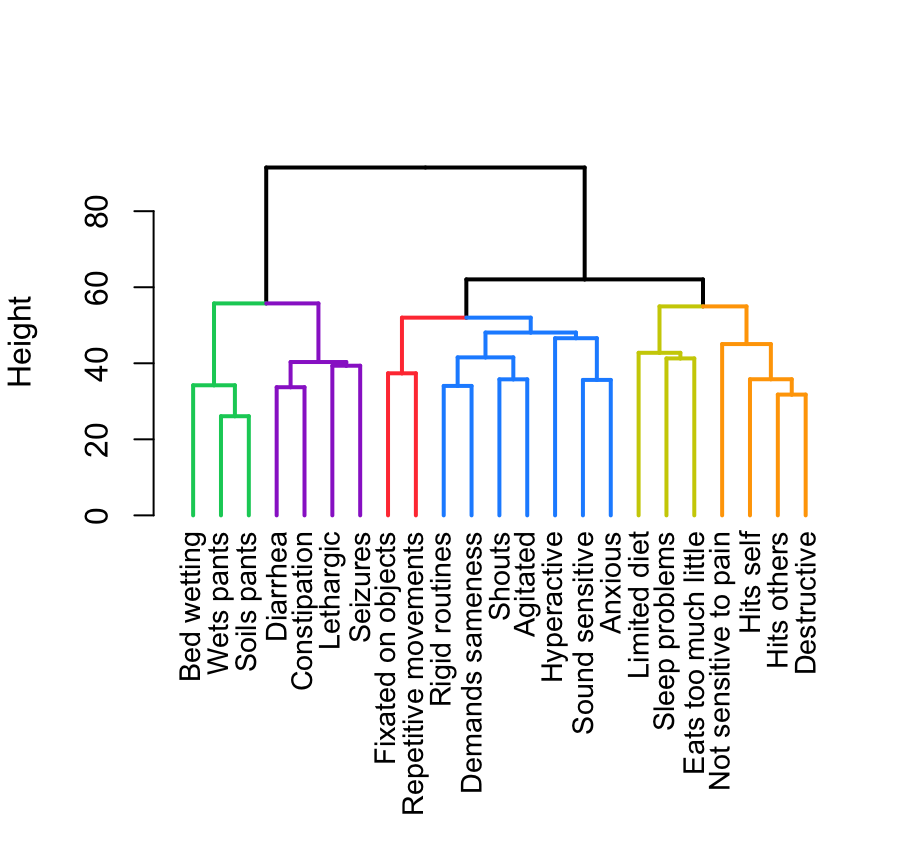

**Figure S18 – Ages 13-22 Dendrogram**

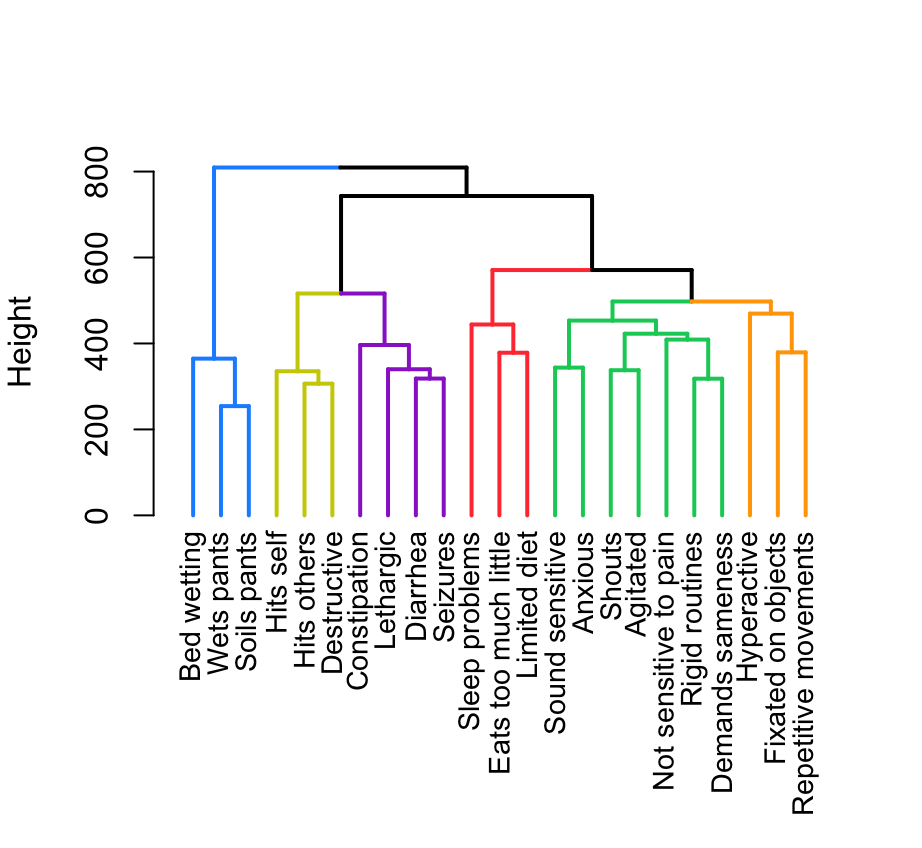

**Figure S19 – Age 2 PCA**

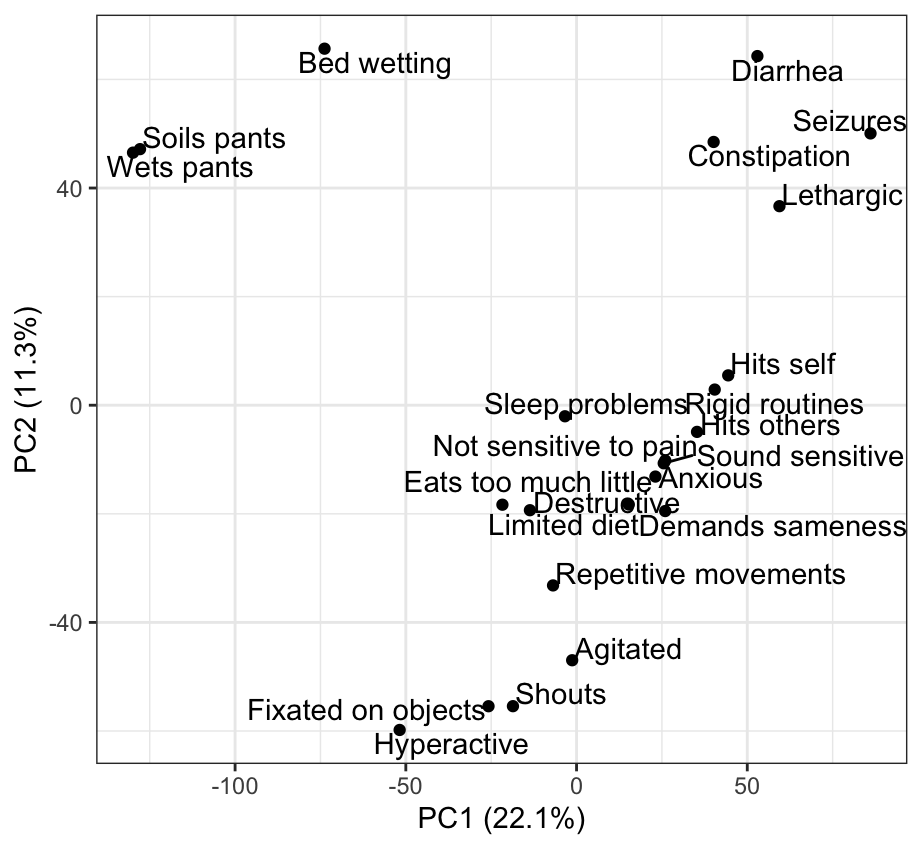

**Figure S20 - Age 3 PCA**

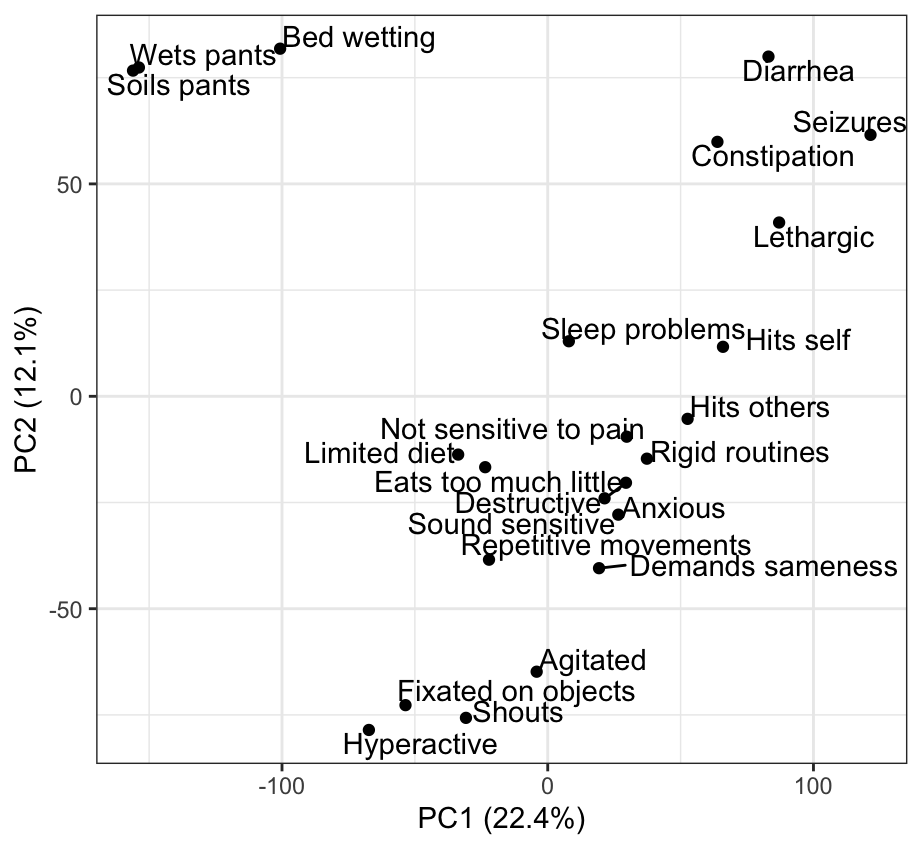

**Figure S21 - Age 4 PCA**

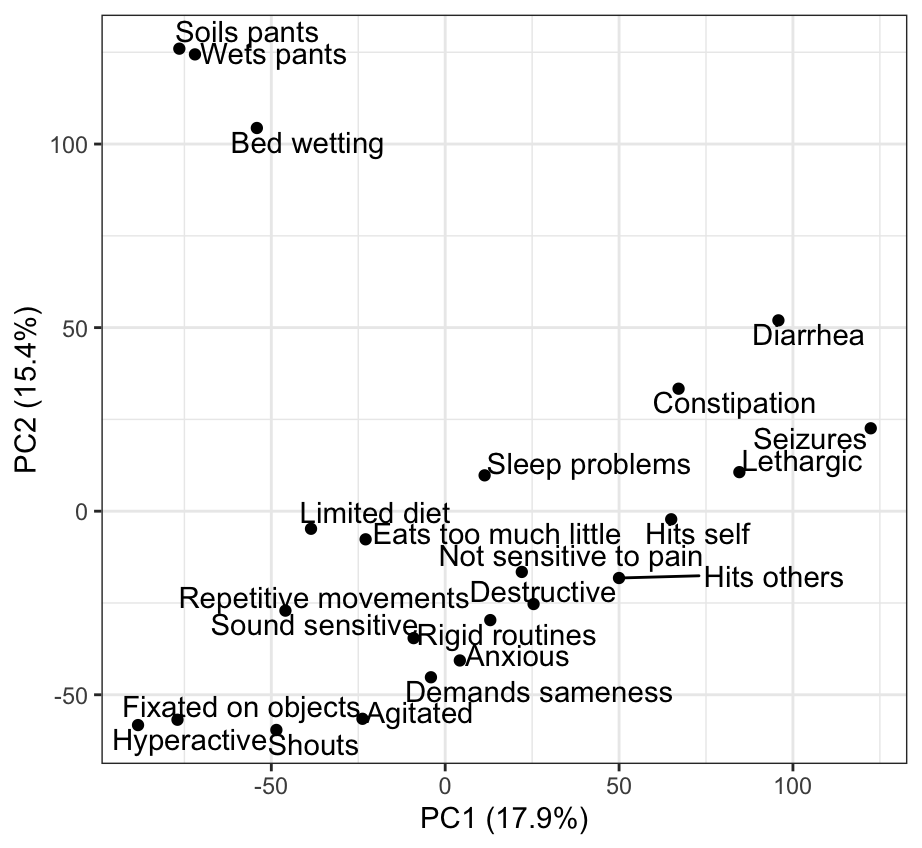

**Figure S22 - Age 5 PCA**

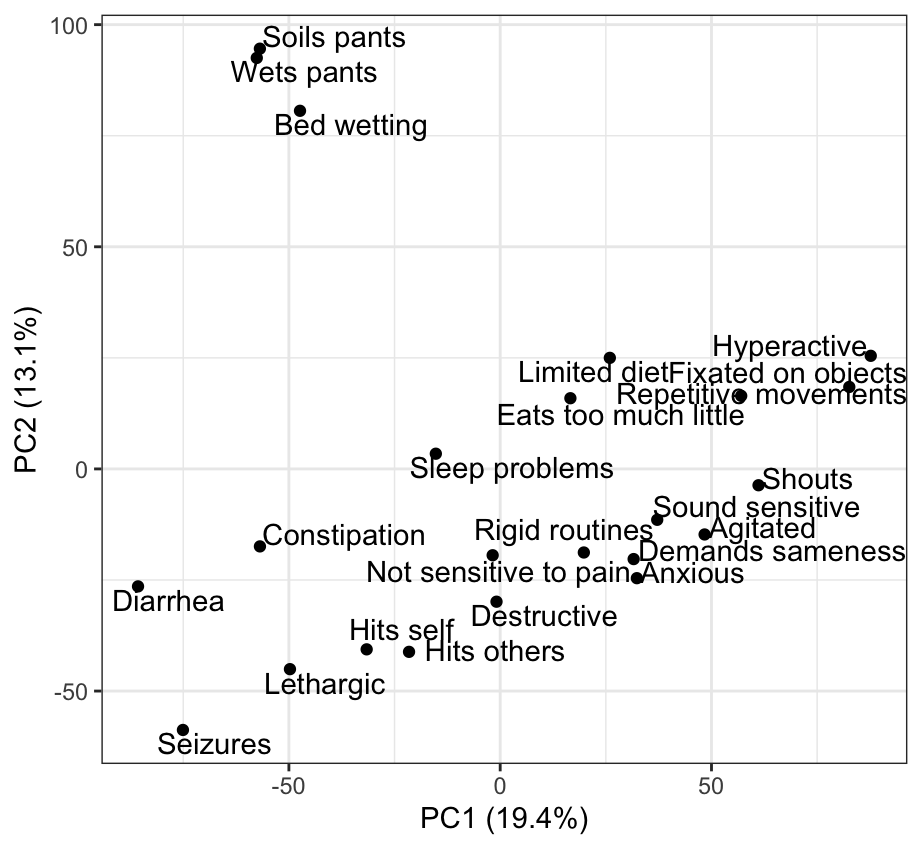

**Figure S23 - Age 6 PCA**

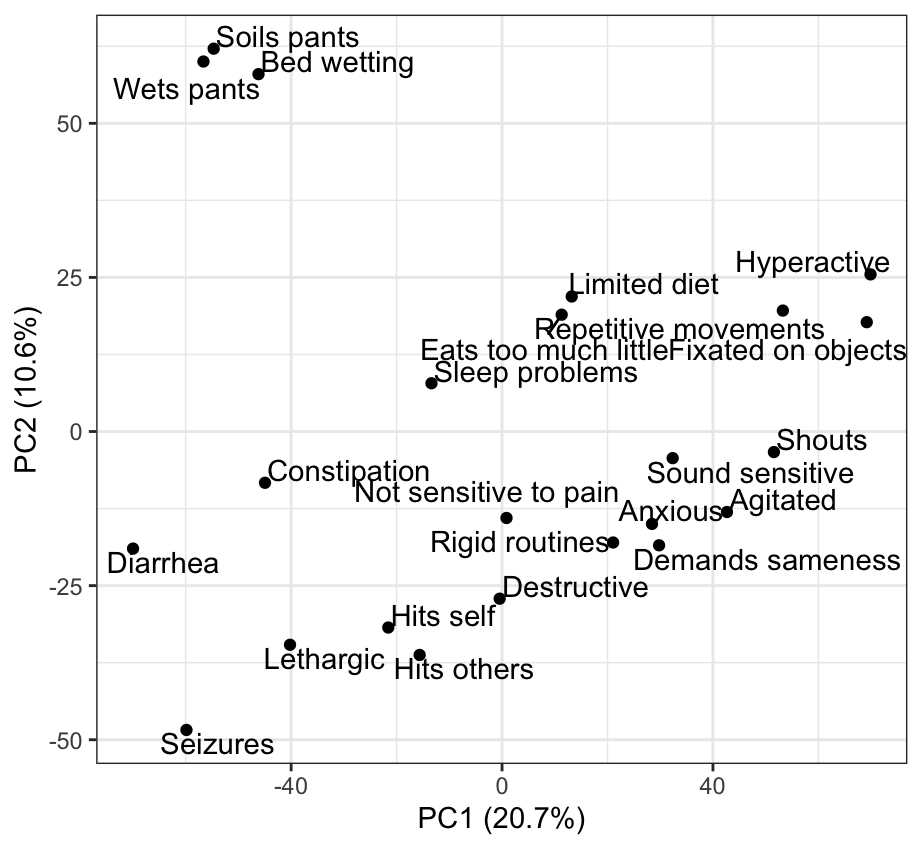

**Figure S24 - Age 7 PCA**

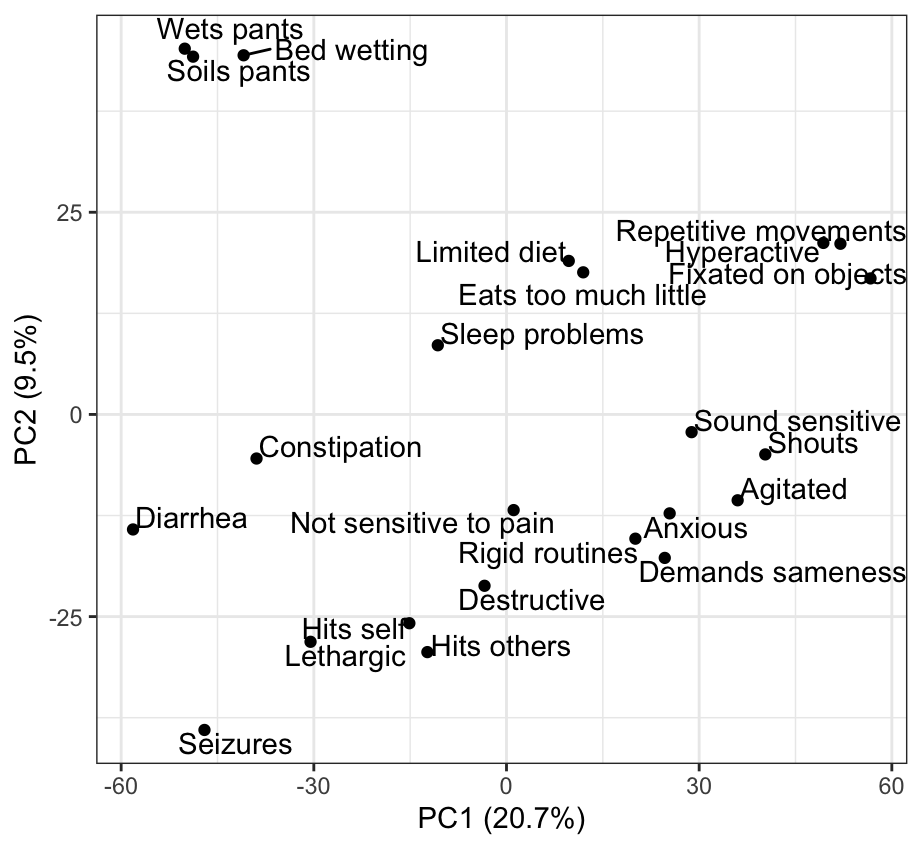

**Figure S25 - Age 8 PCA**

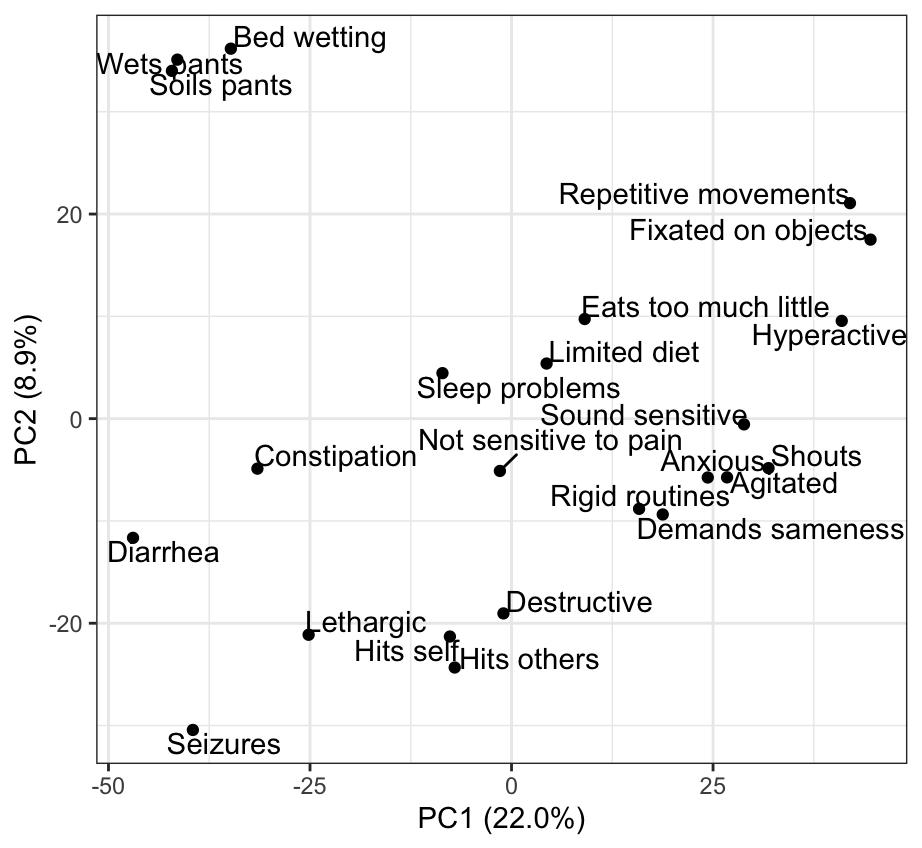

**Figure S26 - Age 9 PCA**

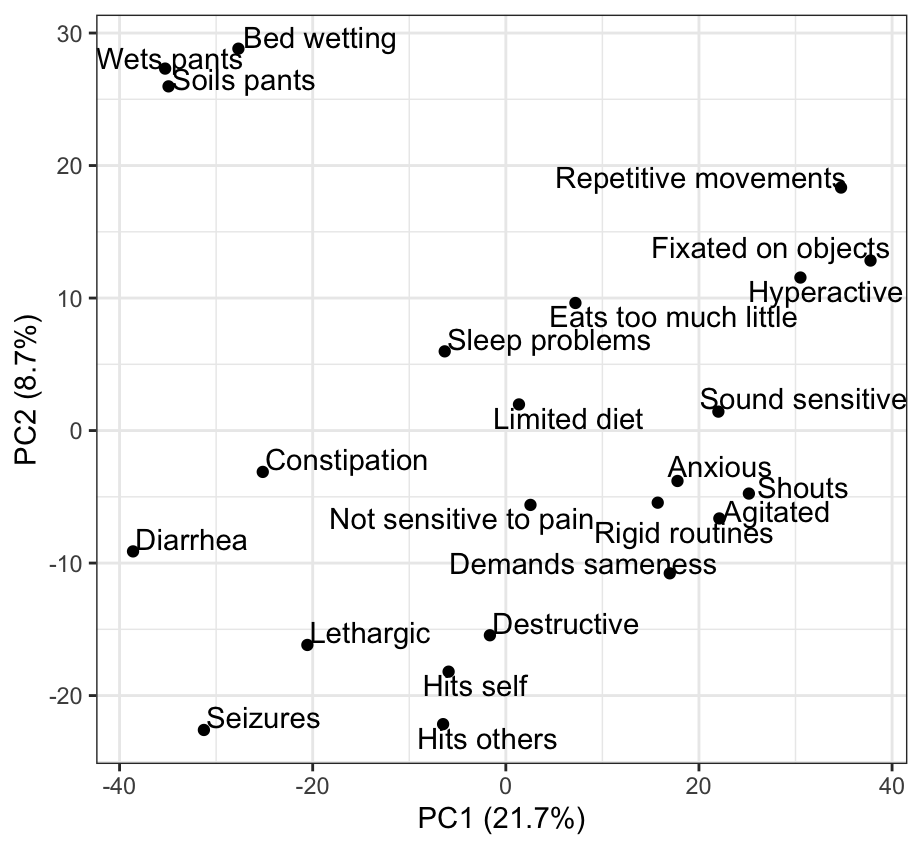

**Figure S27 - Age 10 PCA**

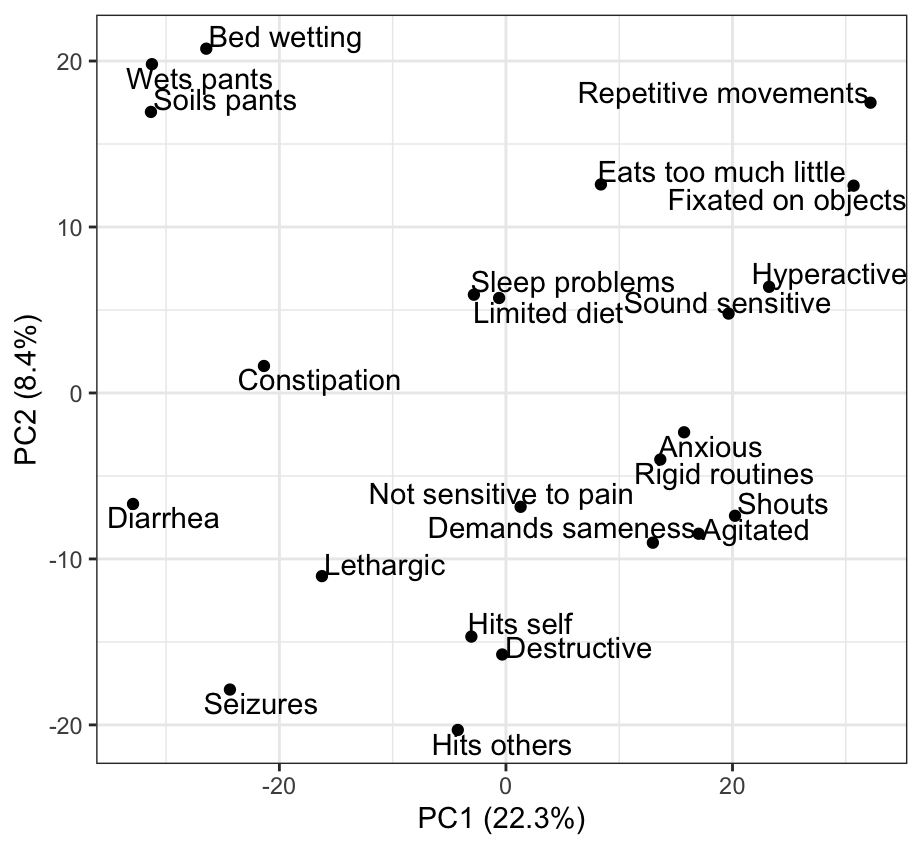

**Figure S28 - Age 11 PCA**

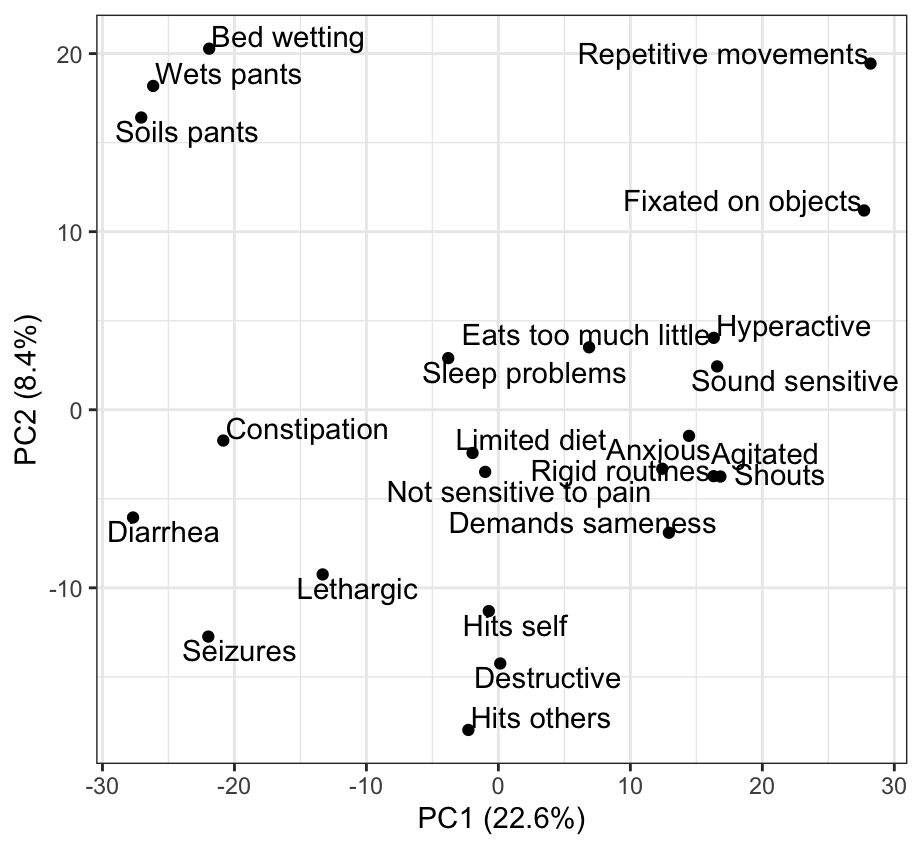

**Figure S29 - Age 12 PCA**

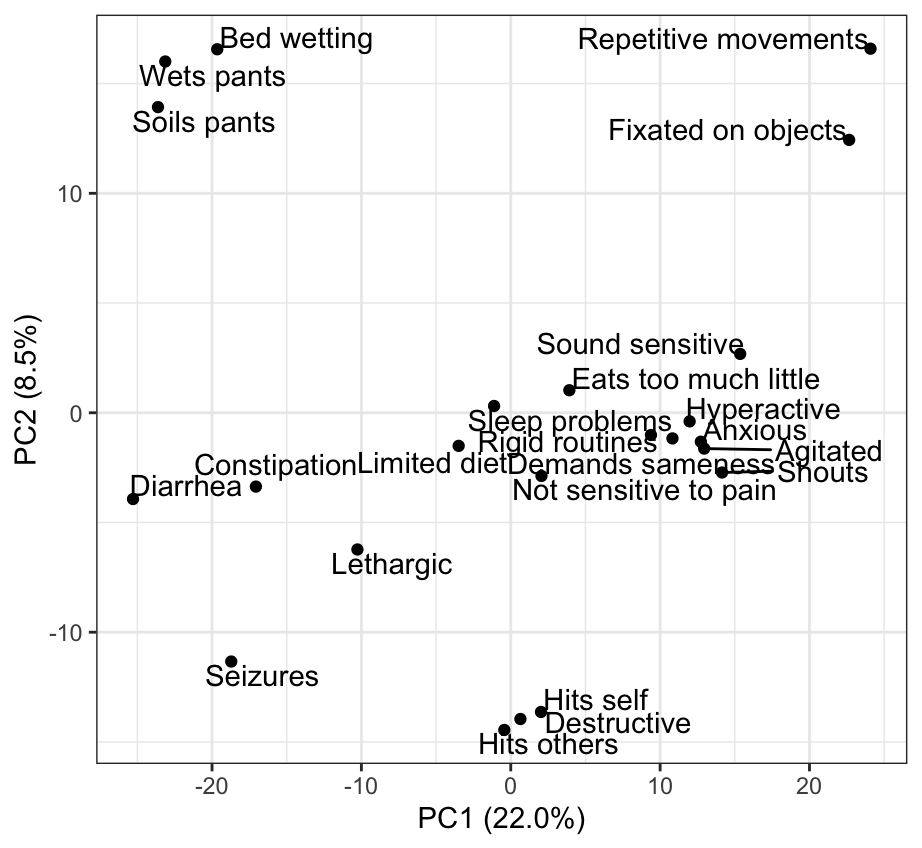

**Figure S30 – Ages 13-22 PCA**

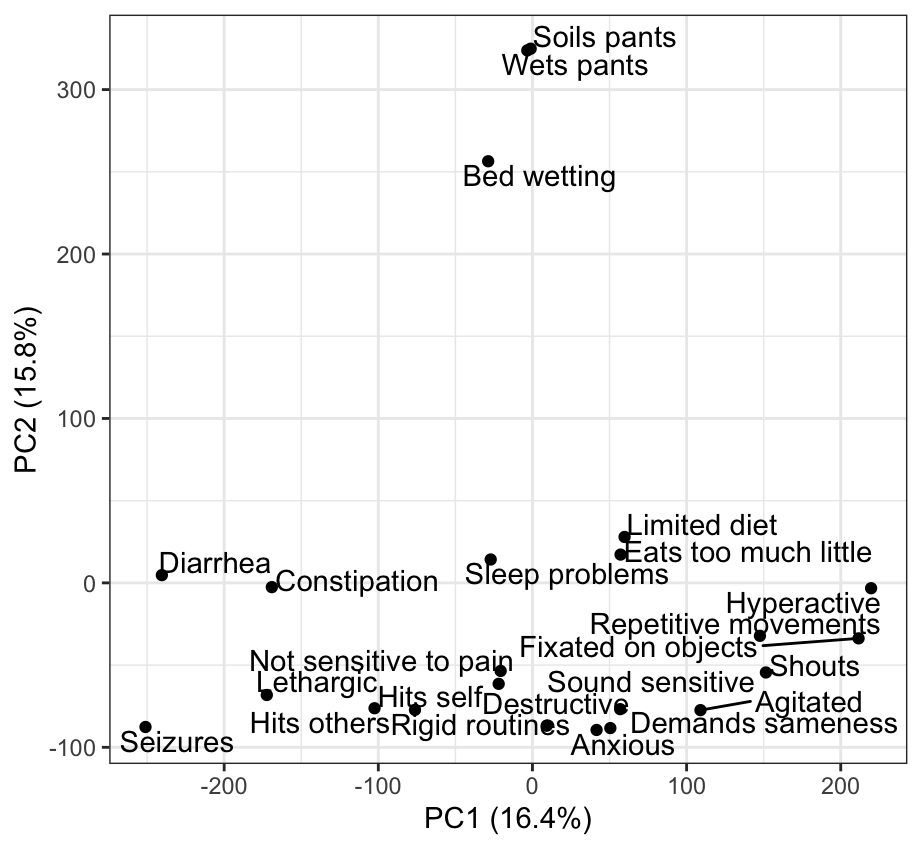

**Figure S31 - Age 13 Dendrogram**

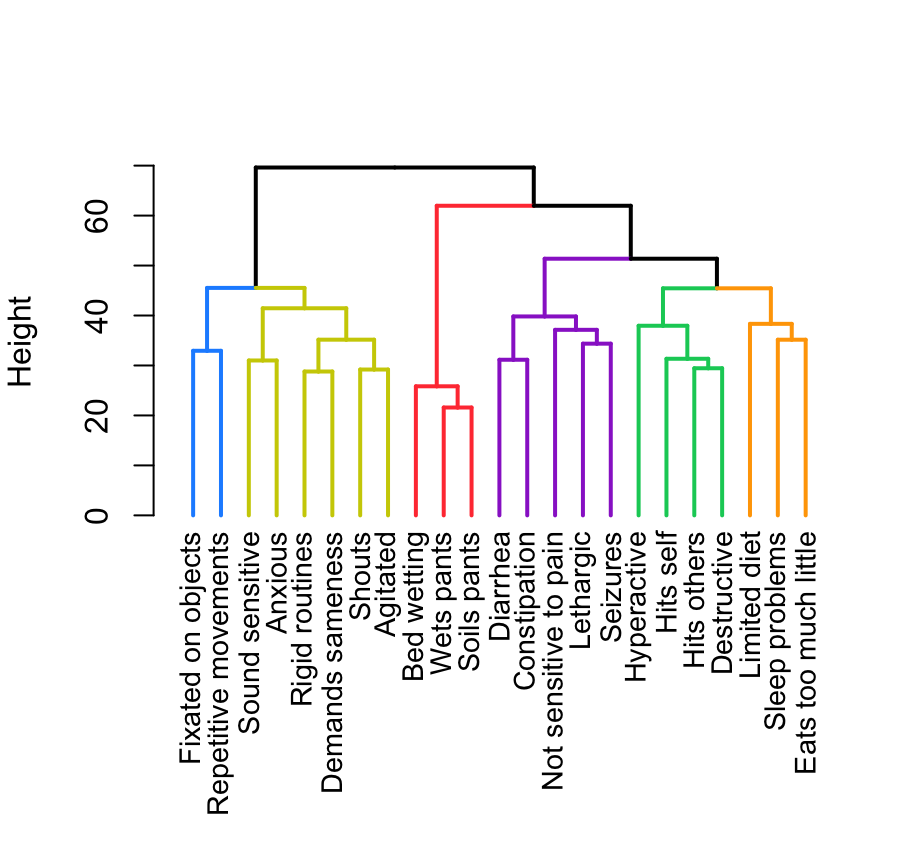

**Figure S32 - Age 14 Dendrogram**

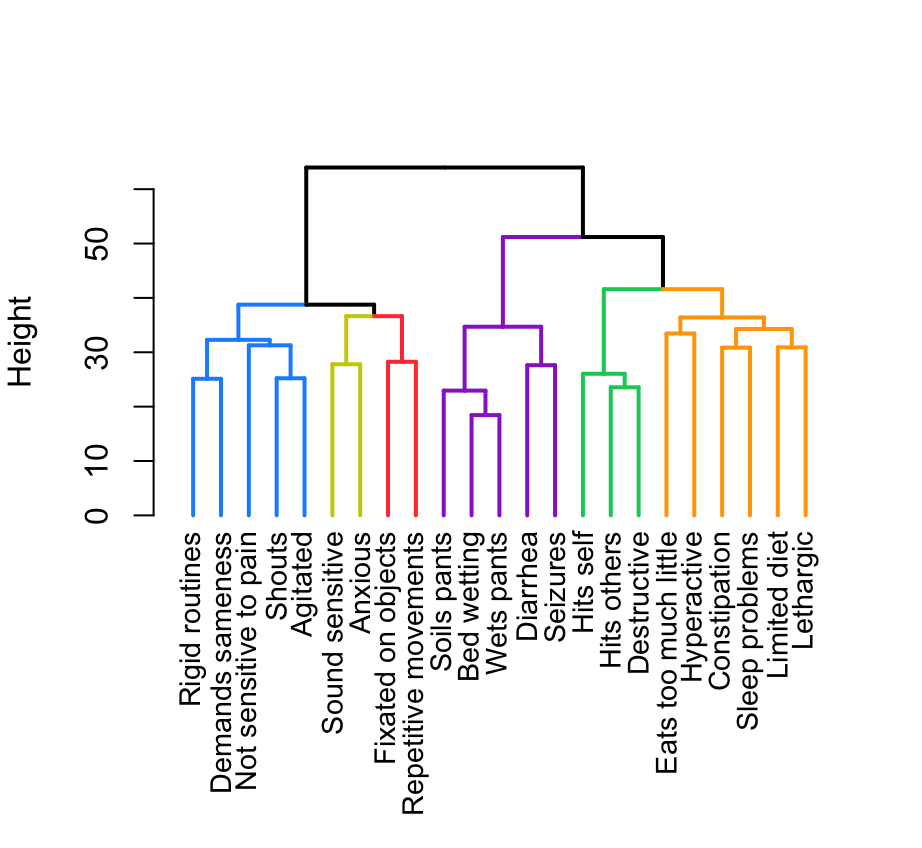

**Figure S33 - Age 15 Dendrogram**

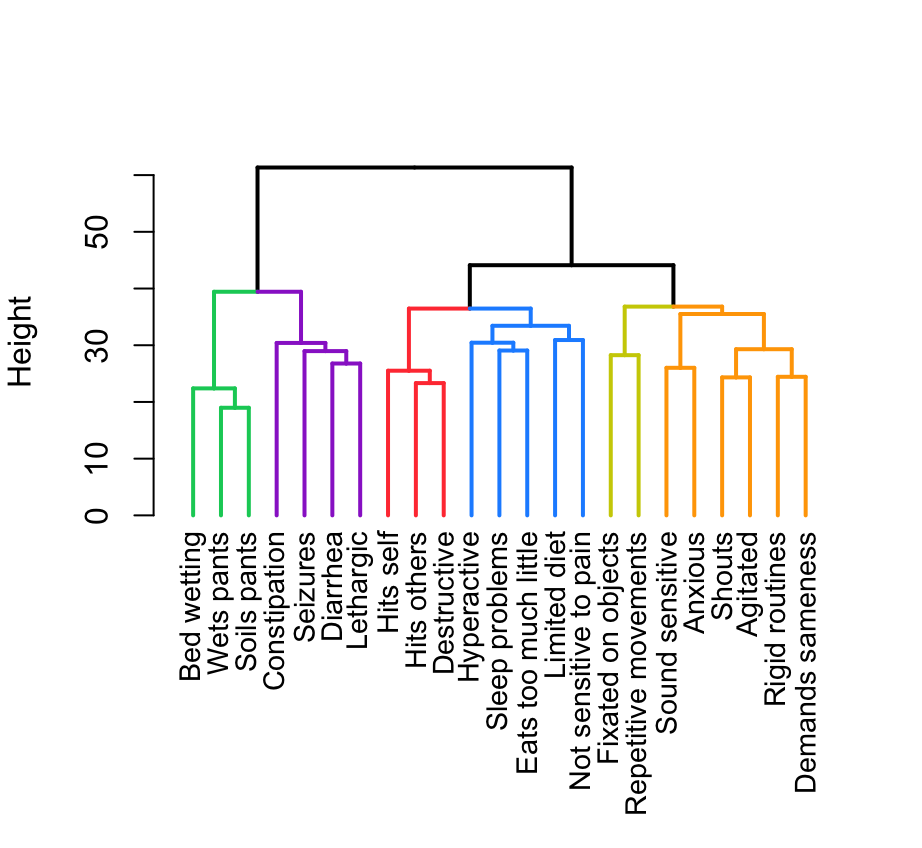

**Figure S34 - Age 16 Dendrogram**

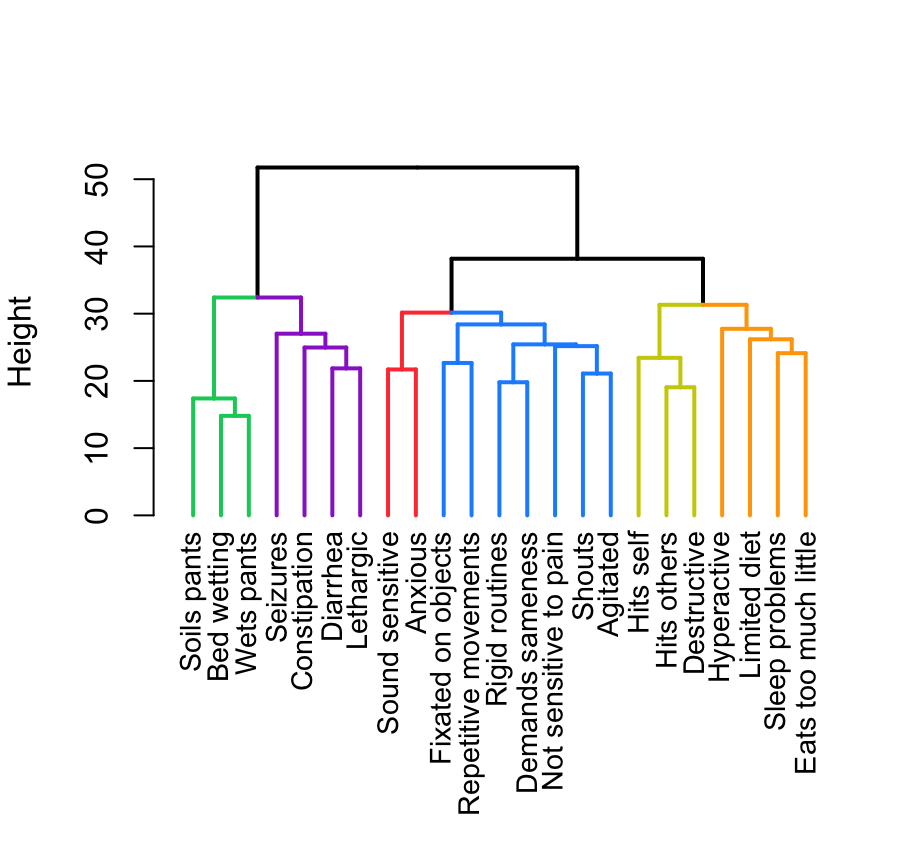

**Figure S35 - Age 17 Dendrogram**

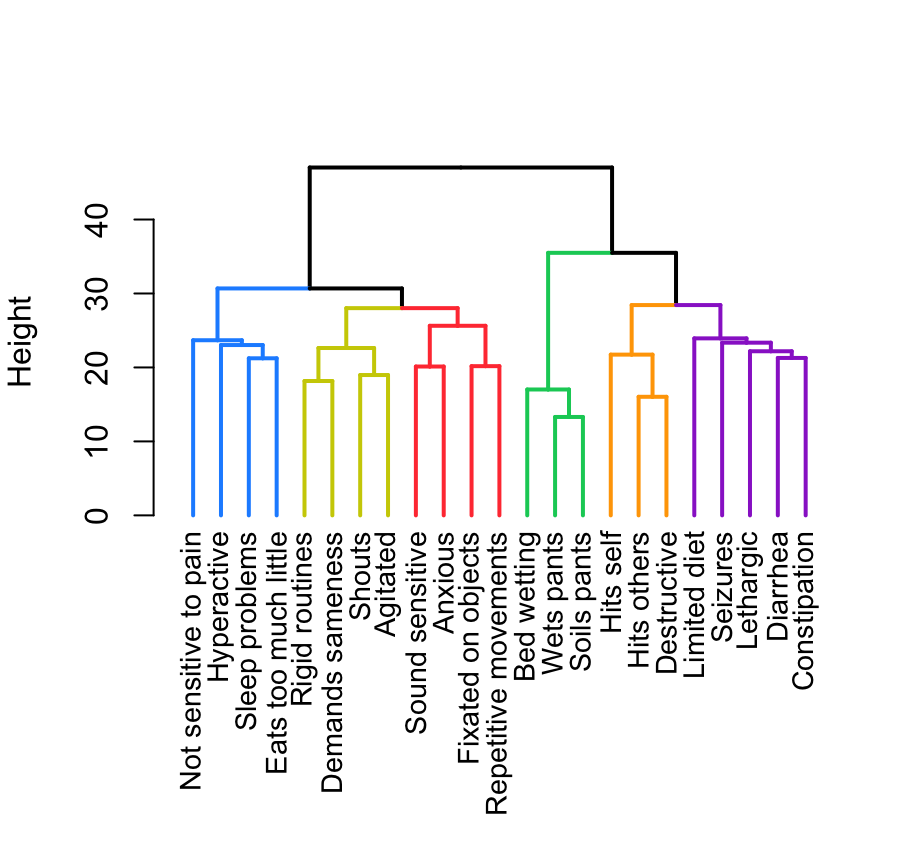

**Figure S36 - Age 18 Dendrogram**

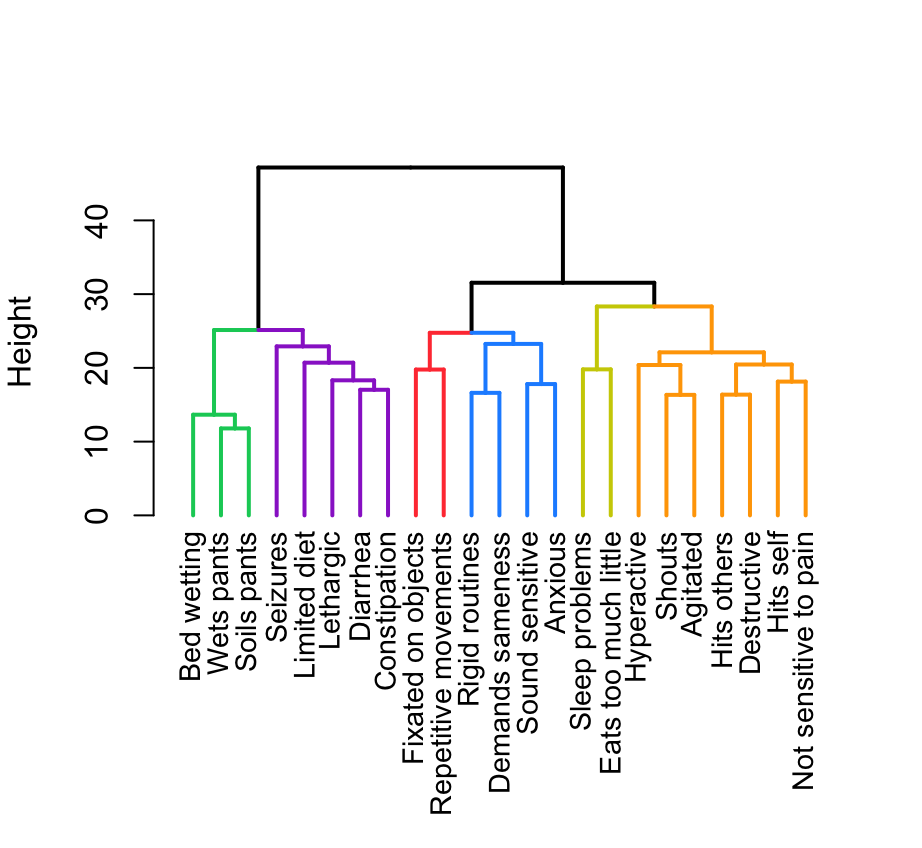

**Figure S37 - Age 19 Dendrogram**

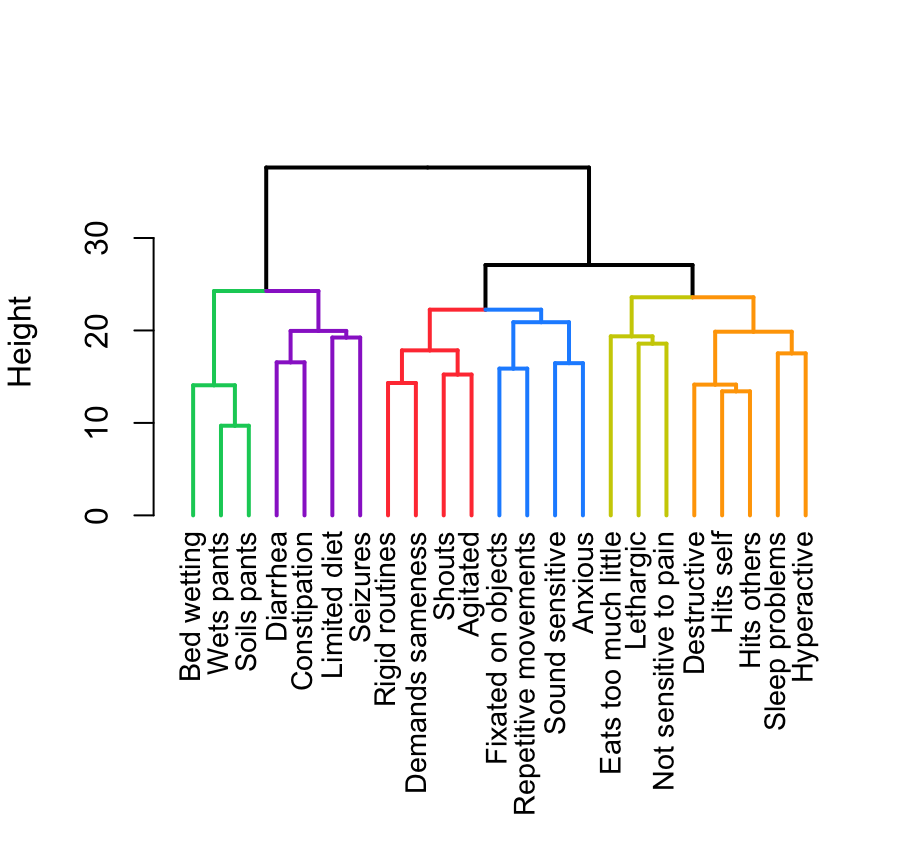

**Figure S38 - Age 20 Dendrogram**

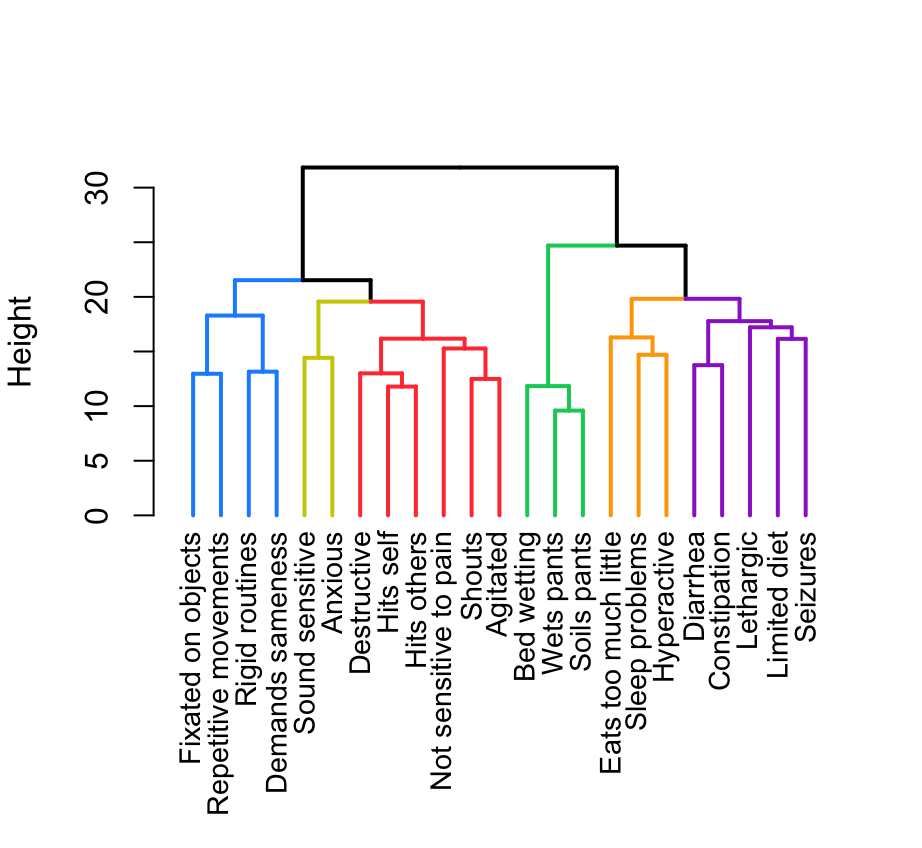

**Figure S39 - Age 21 Dendrogram**

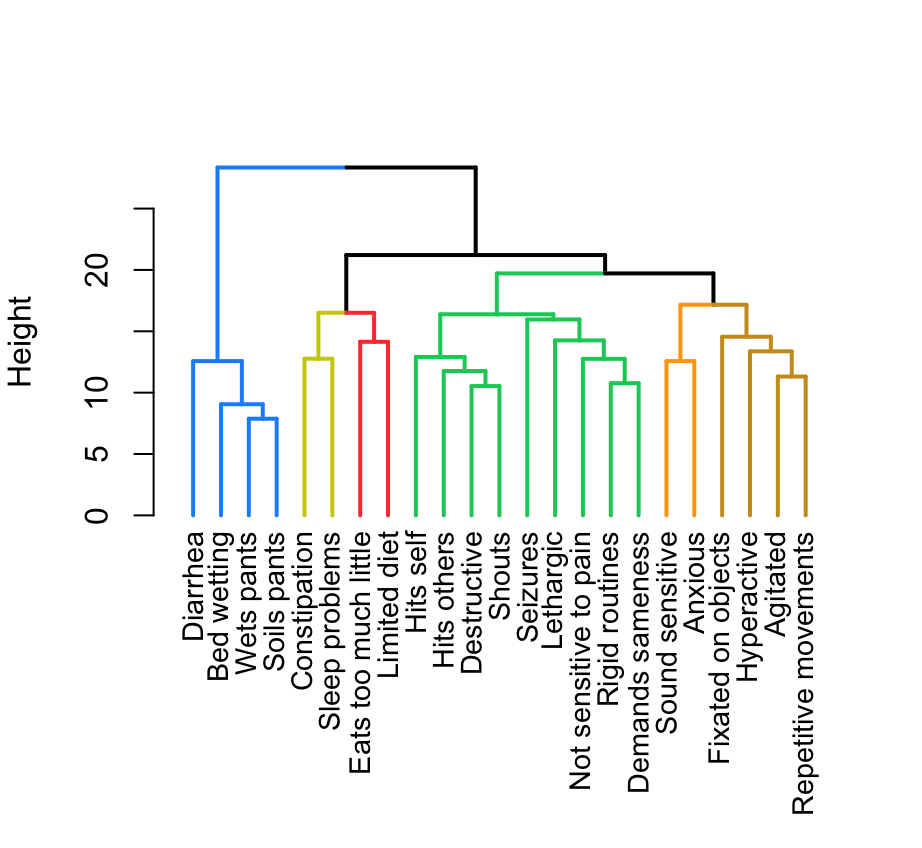

**Figure S40 - Age 22 Dendrogram**

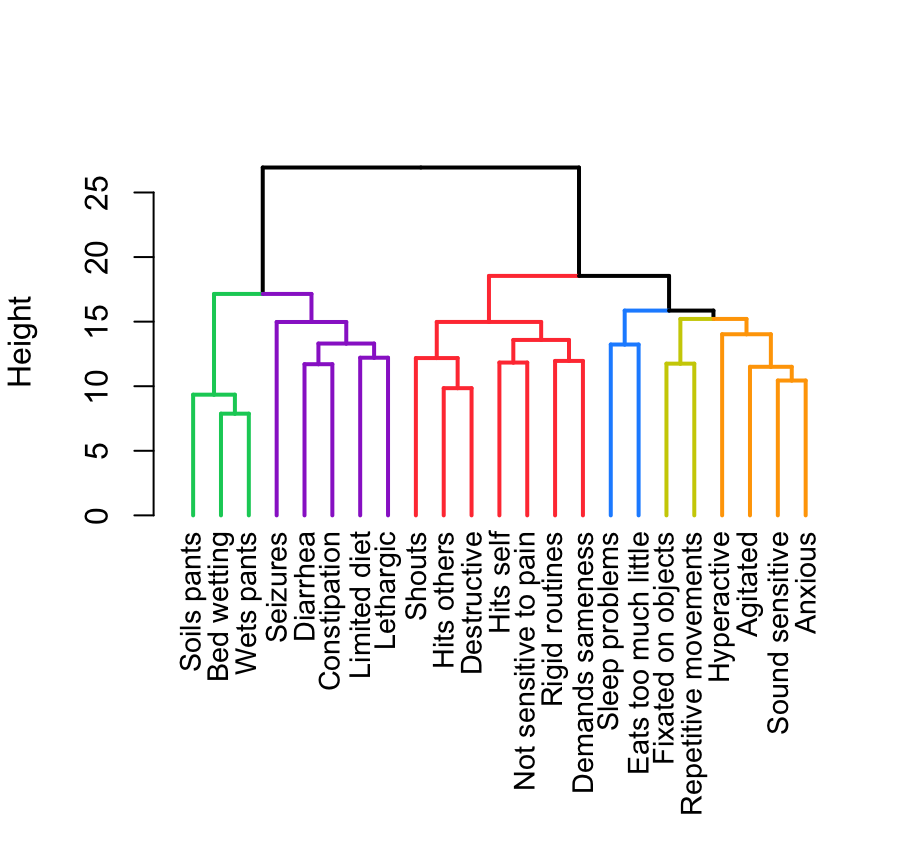

**Figure S41 - Age 13 PCA**

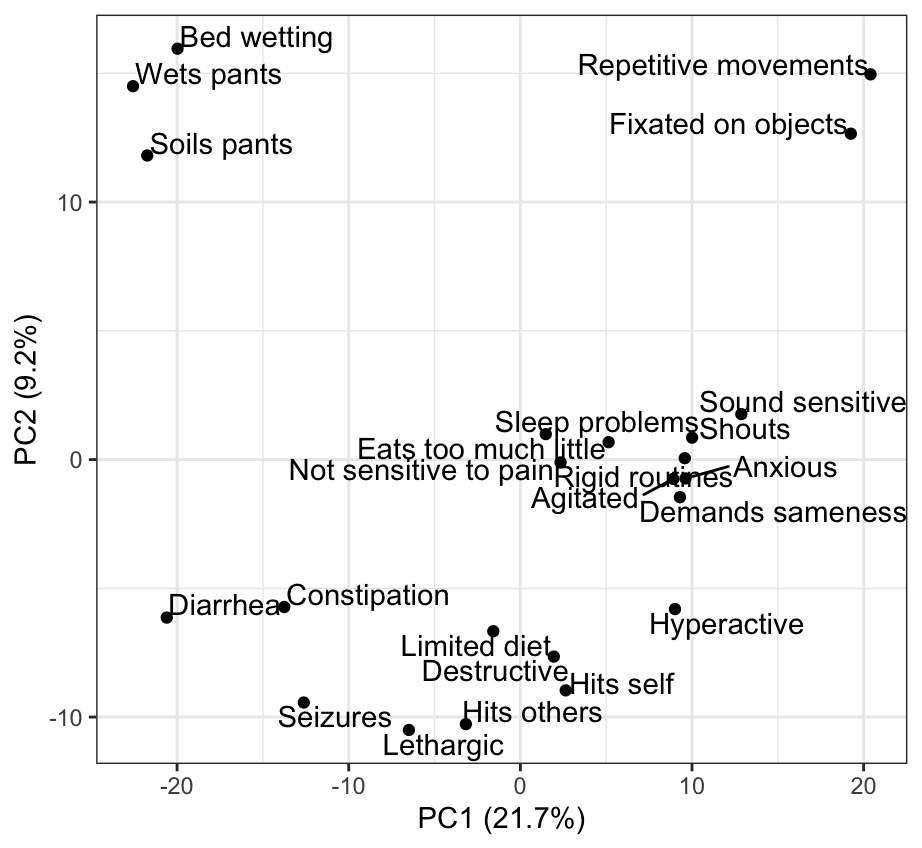

**Figure S42 - Age 14 PCA**

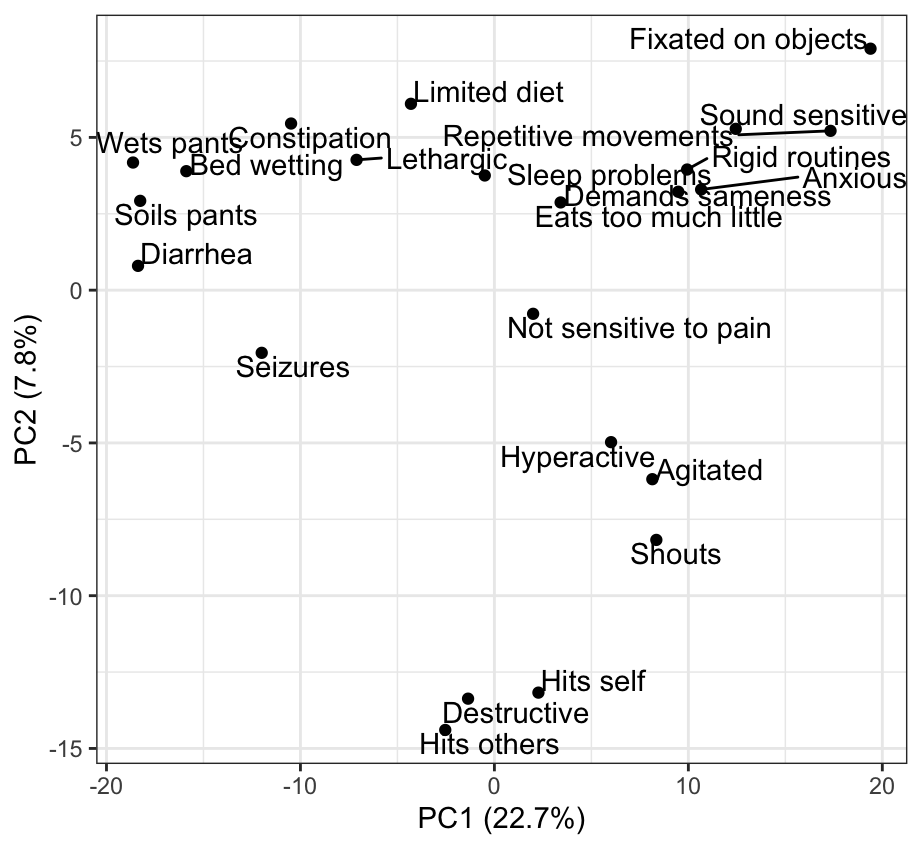

**Figure S43 - Age 15 PCA**

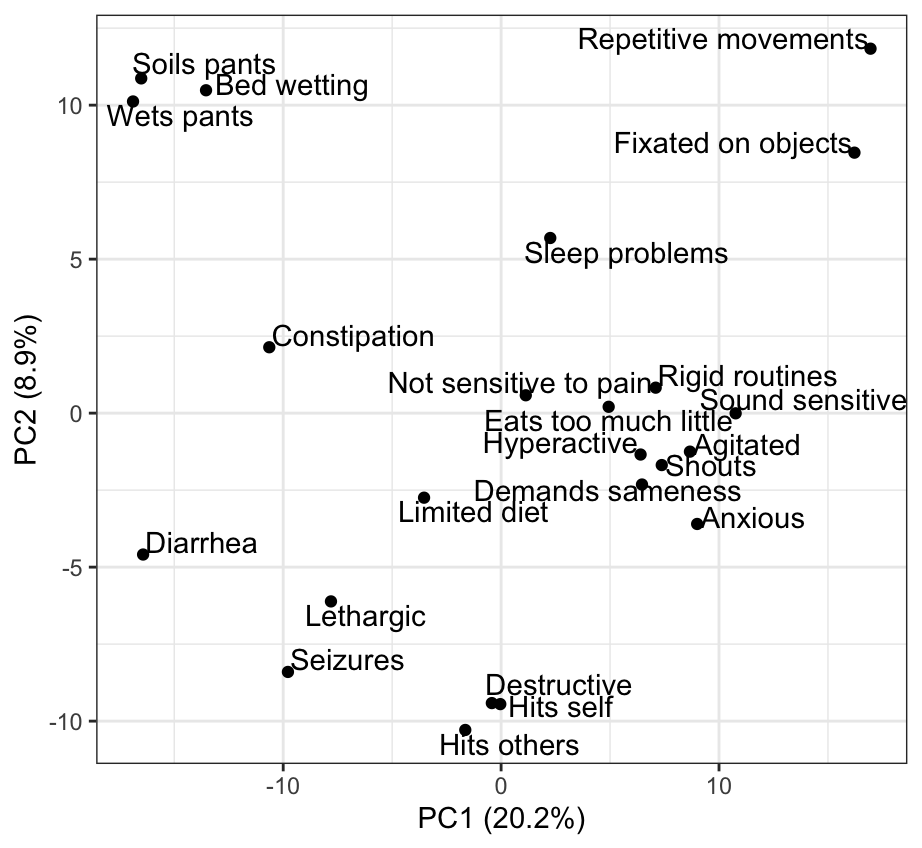

**Figure S44 - Age 16 PCA**

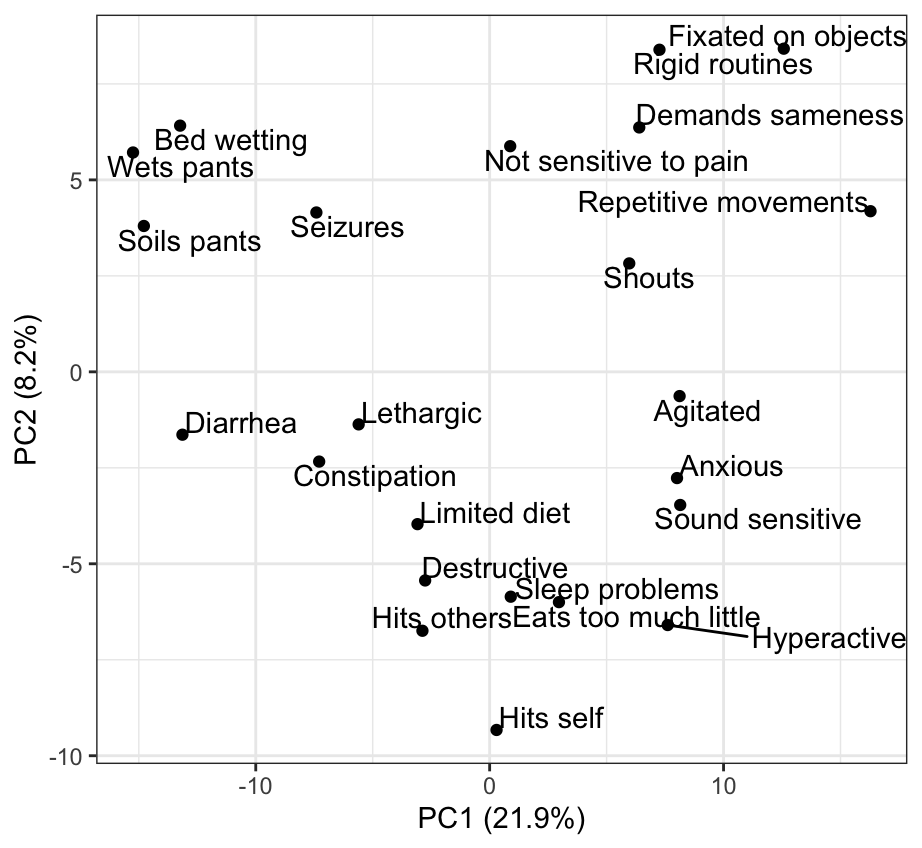

**Figure S45 - Age 17 PCA**

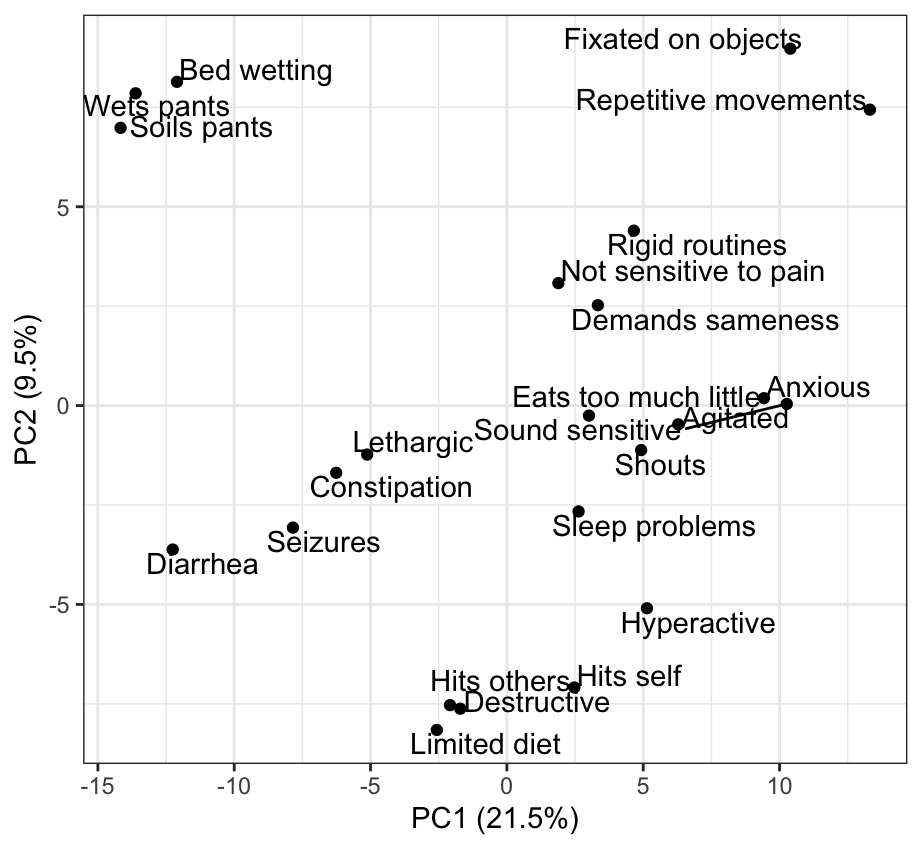

**Figure S46 - Age 18 PCA**

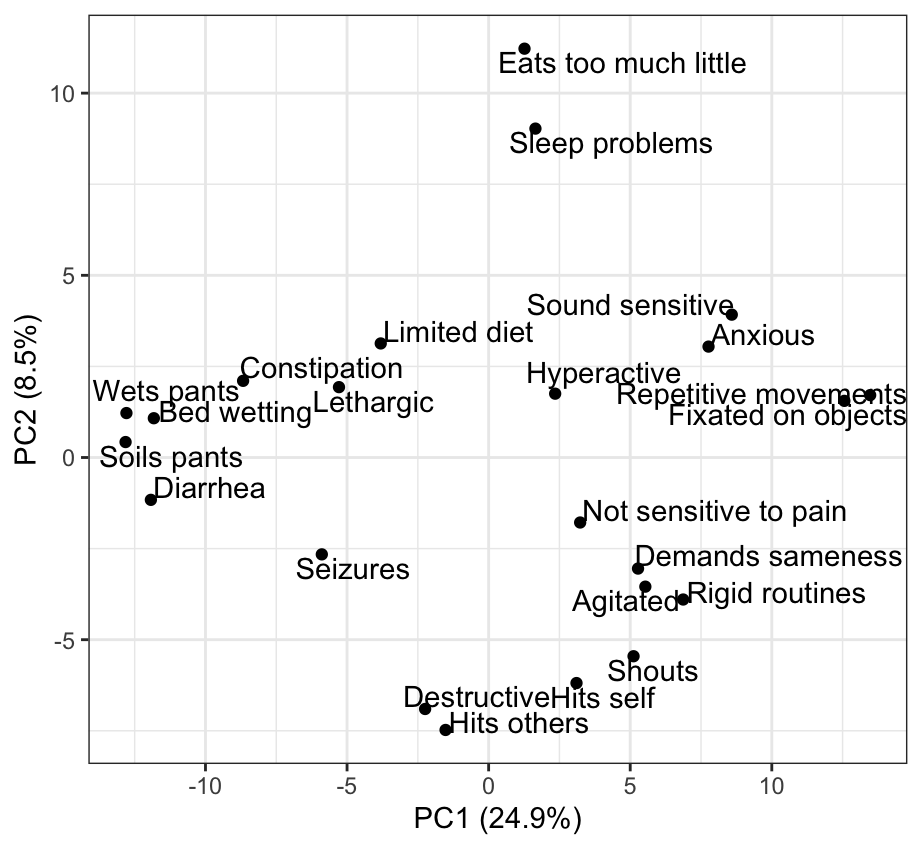

**Figure S47 - Age 19 PCA**

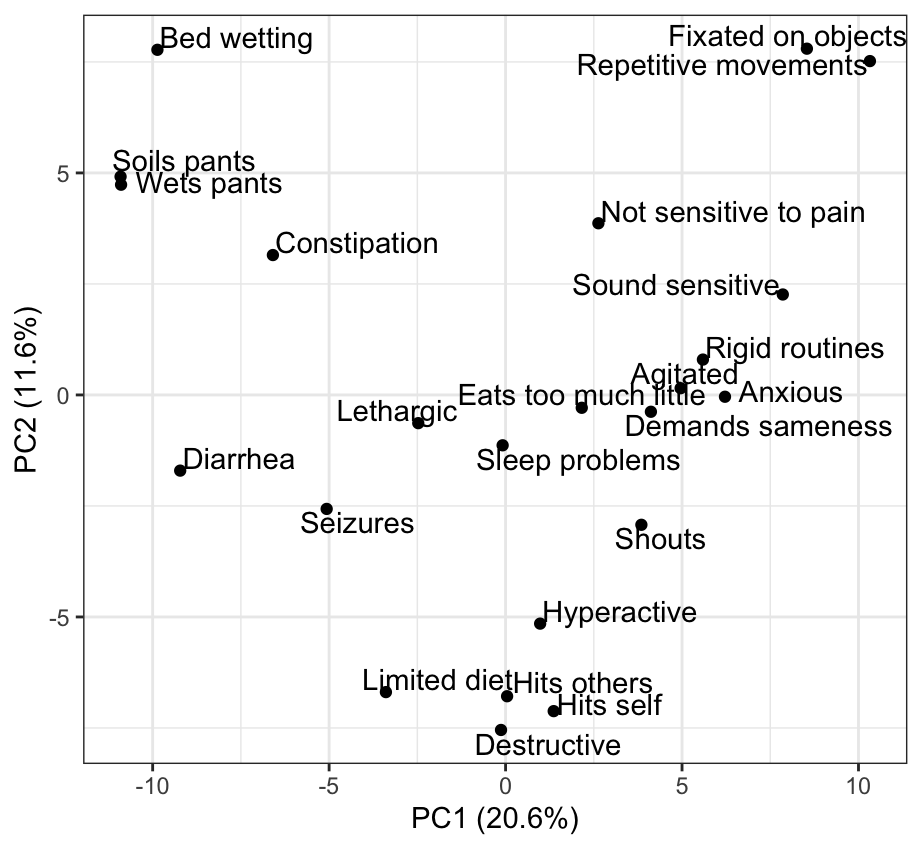

**Figure S48 - Age 20 PCA**

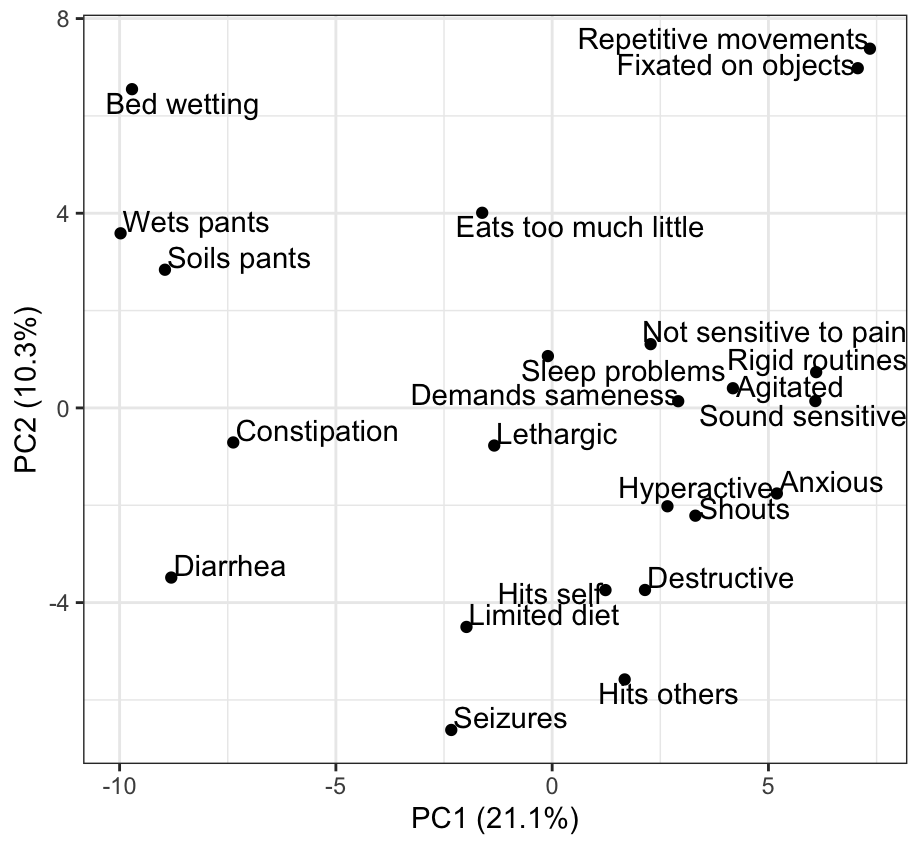

**Figure S49 - Age 21 PCA**

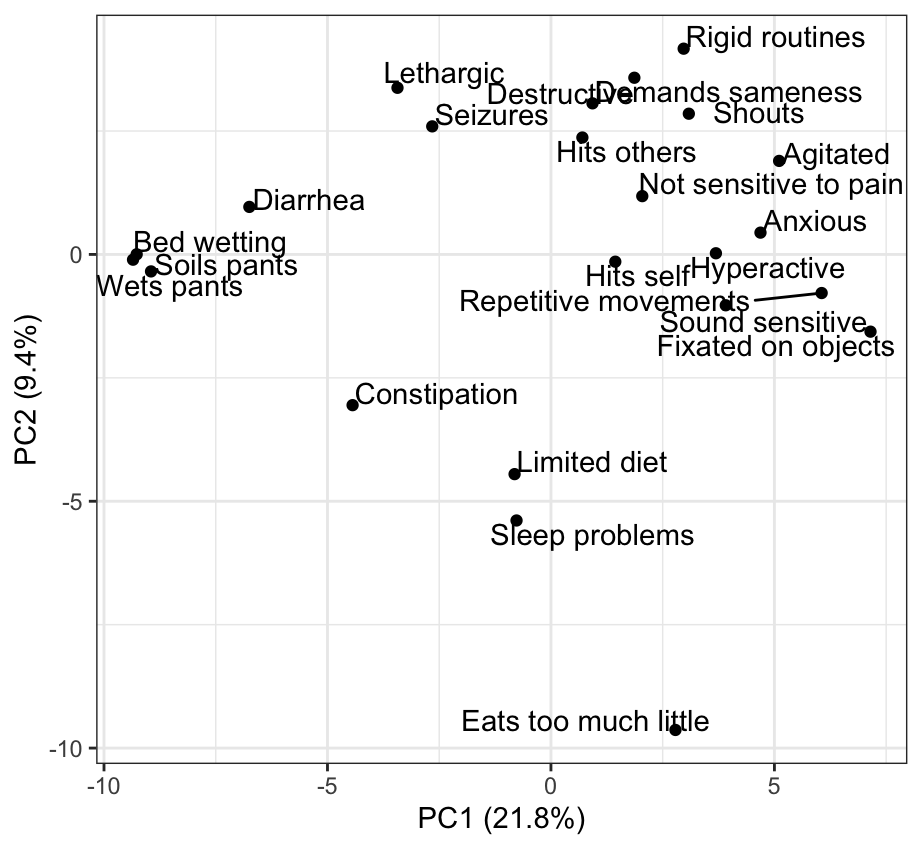

**Figure S50 - Age 22 PCA**

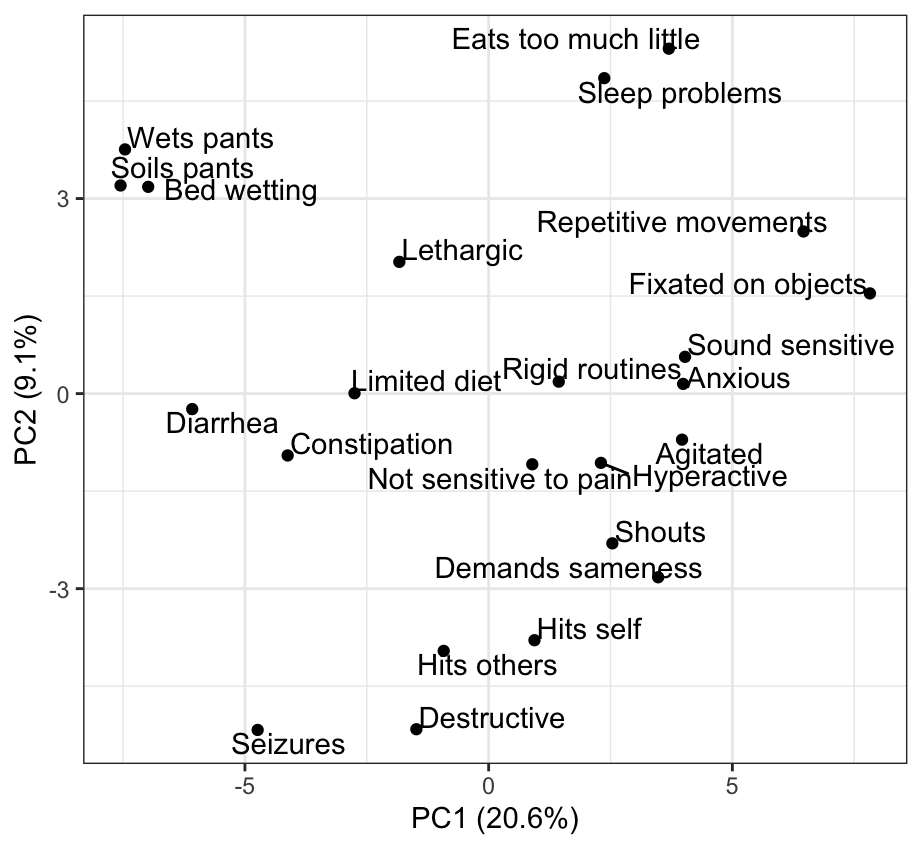

**Table S1 – Results of Principal Component Analysis by Age**

| Age | PC1 | PC2 | Total Variance |
| --- | --- | --- | --- |
| 2 | 22.1% | 11.3% | 33.4% |
| 3 | 22.4% | 12.1% | 34.5% |
| 4 | 17.9% | 15.4% | 33.3% |
| 5 | 19.4% | 13.1% | 32.5% |
| 6 | 20.7% | 10.6% | 31.3% |
| 7 | 20.7% | 9.5% | 30.2% |
| 8 | 22.0% | 8.9% | 30.9% |
| 9 | 21.7% | 8.7% | 30.4% |
| 10 | 22.3% | 8.4% | 30.7% |
| 11 | 22.6% | 8.4% | 31.0% |
| 12 | 22.0% | 8.5% | 30.5% |
| 13-22 | 16.4% | 15.8% | 32.2% |
